# EXTENDING THE SUSCEPTIBLE-EXPOSED-INFECTED-REMOVED (SEIR) MODEL TO HANDLE THE HIGH *fa*LSE *n*EGATIVE RATE AND *sy*MPTOM-BASED ADMINISTRATION OF COVID-19 DIAGNOSTIC TESTS: *SEIR-fansy*

**DOI:** 10.1101/2020.09.24.20200238

**Authors:** Ritwik Bhaduri, Ritoban Kundu, Soumik Purkayastha, Michael Kleinsasser, Lauren J. Beesley, Bhramar Mukherjee

## Abstract

The false negative rate of the diagnostic RT-PCR test for SARS-CoV-2 has been reported to be substantially high. Due to limited availability of testing, only a non-random subset of the population can get tested. Hence, the reported test counts are subject to a large degree of selection bias. We consider an extension of the Susceptible-Exposed-Infected-Removed (SEIR) model under both selection bias and misclassification. We derive closed form expression for the basic reproduction number under such data anomalies using the next generation matrix method. We conduct extensive simulation studies to quantify the effect of misclassification and selection on the resultant estimation and prediction of future case counts. Finally we apply the methods to reported case-death-recovery count data from India, a nation with more than 5 million cases reported over the last seven months. We show that correcting for misclassification and selection can lead to more accurate prediction of case-counts (and death counts) using the observed data as a beta tester. The model also provides an estimate of undetected infections and thus an undereporting factor. For India, the estimated underreporting factor for cases is around 21 and for deaths is around 6. We develop an R-package *SEIR-fansy* for broader dissemination of the methods.

## 1 Introduction

The COVID-19 pandemic, caused by severe acute respiratory coronavirus 2 (SARS-CoV-2), was first identified in Wuhan, China in December 2019. The World Health Organisation declared the outbreak a Public Health Emergency of International Concern on 30 January 2020. On 11 March, 2020, the outbreak was classified a pandemic. As of August 15, 2020, more than 21 million cases of COVID-19 have been reported in 188 countries and territories. More than 775,000 deaths have been reported worldwide [25].

Ramping up rapid testing for COVID-19 is an important component of proposed non-pharmaceutical intervention strategies to control an outbreak [21]. Presence of an active COVID-19 infection can be confirmed by using reverse transcription polymerase chain reaction (RT-PCR) tests of infected secretions (typically obtained via a nasopharyngial swab) or CT imaging of the chest [1]. Evidence-based clinical understanding of diagnostic test results is essential to garner insight about COVID-19 management through effective contact tracing and isolation, ultimately leading to mitigated transmission risk [27].

There has been growing concern about false negative RT-PCR tests in patients with COVID-19-like symptoms. A preprint [31] describes a study of 213 patients hospitalized with COVID-19. In the week following onset of disease for the patients, 11% of sputum-, 27% of nasal- and 40% of throat-based samples were deemed as false negatives. Studying publicly available time-series data of laboratory tests of SARS-CoV-2 viral infection, another article [6] aims to better understand misclassification errors in the identification of true COVID-19 cases using Bayesian methods. A systematic review [2] of five studies (enrolling a total of 957 patients) reports a range of 2 to 29% of false negatives. The authors note that the certainty of evidence is suspect, because of the heterogeneity of sensitivity estimates among the studies, lack of blinding to index test results in establishing diagnoses, and failure to report key RT-PCR characteristics. The general consensus on the issue of false negatives is that the evidence, while limited, raises serious concerns about negative RT-PCR results. In particular, false negative results are worrisome because they might provide false assurance to someone with a true infection and lead to further spread of the disease [29].

Models for projecting infectious disease spread have become widely popular in the wake of the pandemic. Some popular models include the ones developed at the Institute of Health Metrics (IHME)[7] (University of Washington, Seattle) and at the Imperial College London (ICL)[9]. The IHME COVID-19 project initially relied on an extendable nonlinear mixed effects model for fitting parametrized curves to COVID-data, before moving to a compartmental model to analyze the pandemic and generate projections. The ICL model calculates backwards from observed deaths to estimate transmission that occurred several weeks previously, allowing for the time lag between infection and death. A Bayesian mechanistic model is introduced - linking the infection cycle to observed deaths, inferring the total population infected (attack rates) as well as the time-varying reproduction number *R*(*t*) - an important public health metric. With the onset of the pandemic, there has been renewed interest in multi-compartment models, which have played a central role in modeling infectious disease dynamics since the 20th century [23]. The simplest of compartmental models include the standard SIR [13] model, which has been extended [22] to incorporate various types of time-varying quarantine protocols, including government-level macro isolation policies and community-level micro inspection measures. Further extensions include one which adds a spatial component to this temporal model by making use of a cellular automata structure [32]. Larger SEIR models incorporate different states of transition between susceptible, exposed, infected and removed compartments and have been used in the early days of the pandemic in the Wuhan province of China [30]. The SEIR model has been further extended to the SAPPHIRE model [12], which accounts for the infectiousness of asymptomatic [4] and presymptomatic [24] individuals in the population (both of which are crucial transmission features of COVID-19), time varying ascertainment rates, transmission rates and population movement. All these models use the daily time series of susceptible, infected, recovered (and sometimes fatalities) to model the full course of disease transmission. None of the models mentioned above address the potential issue of false negatives, which also contribute to the unreported cases (along with those untested infectious cases).

An auxiliary concern surrounding testing is selection bias, introduced by the selection of individuals (or sub-groups) that are prioritized for receiving testing. The testing guidelines are often driven by severity of symptoms. Sometimes tests are offered based on occupation, as a routine check prior to undergoing any medical procedure, or based on pre-existing health conditions. The sample tested is, therefore, not representative of the population intended to be analyzed, and predicted case-counts and estimated parameters can deviate from the truth. While it may be possible to estimate the underlying selection model or run sensitivity analyses, it is difficult to prove that bias has been reduced or eliminated [10, 5]. In the context of COVID-19 testing, such biases will continue to impact estimates of disease prevalence and the effective reproduction number until a random sample of the population is tested and/or testing becomes abundantly available. Moreover, false positive/negative rates of tests interact with selection bias in complex ways. A preprint [8] discusses such interaction in greater detail, noting that current statistical models do not simultaneously account for selection bias and measurement error. The author also puts forth several suggestions on how to improve current case-count reporting after addressing these two issues.

In this paper, we first propose a Bayesian compartmental epidemiological model which accounts for false negatives in RT-PCR tests assuming a known value of the test sensitivity. We introduce compartments for false negatives and untested individuals into the structure of the SEIR model. A system of differential equations connecting each compartment is presented, and we derive expressions for estimating the basic reproduction number *R*_0_ [11] after accounting for imperfect tests. The model provides an estimate for total number of cases and deaths (both reported and unreported) after accounting for the false negatives. Uncertainty estimates for each quantity of interest are provided via Markov chain Monte Carlo draws from the posterior distribution of the model parameters. Simulation studies are used to assess the accuracy of estimation and prediction. We note that though estimates of *R*_0_ remains relatively robust to various choices of FN (False Negative probability of the RT-PCR tests), modeling daily new cases, recoveries, and deaths simultaneously after accounting for FN when it truly exists leads to better estimates of the current number of active cases. From a public health perspective, this quantity is more important than estimating total number of cases as this is directly related to health care needs on a daily basis. Further, we attempt to quantify the effect of selection where tests are offered based on symptoms. Finally, we illustrate our methods by analyzing the transmission patterns of COVID-19 in India to provide national estimates/projections along with state-level estimates for the Indian states of Maharashtra and Delhi, two hotspots for the outbreak. While our method is not country or state specific, we have chosen to analyze the data from India for two main reasons - firstly, the number of daily new cases in India has been steadily increasing from March, and the virus curve has not turned the corner even in August. With 1.38 billion susceptible individuals, India now stands 3^rd^ in the world in terms of the total number of cases and is contributing the highest number of cases and deaths to the Global tally in August. The second reason is that the stages of national lockdowns in India are well defined and the public health policies were roughly uniform throughout the country, which align with the assumptions in our model.

The rest of the paper is organized as follows. In Section 2, we introduce the basic notations and compartments in our model followed by the assumed transmission dynamics. We consider two likelihoods: (1) a binomial/Poisson likelihood using only the data on daily cases and (2) a multinomial likelihood where we jointly model daily cases, recoveries, and deaths. We describe estimation strategies and uncertainty quantification of model estimates. We provide an R-package *SEIR-fansy*(*fa*LSE *n*EGATIVE rate and *sy*MPTOM) to implement our model. In Section 3, we present extensions of this core model structure to incorporate (i) time varying fatality rates, changing through the course of the pandemic, (ii) symptom-based testing of the infected and (iii) the selection bias related to who gets tested. Section 4 contains analysis of the data from India. In Section 5, we conduct an extensive simulation study to assess the performance of our model under misclassification and symptom-related selection. We also use simulations to characterize the influence of increasing the number of available tests and its impact on the peak and duration of the pandemic. Section 6 provides concluding discussion.

## 2 Methods and Notation

Compartmental models are mathematical vehicles to study the spread of an infectious disease. At a given time, members of the population fall into various model compartments based on their current status (e.g. Susceptible-**S**, Exposed-**E**, Infectious-**I**, or Recovered-**R** for the SEIR structure). The model structure describes the flow patterns of individuals between compartments over time. In this framework, temporal dynamics of the disease process can be viewed as continuous or occurring in discrete time intervals. Many compartmental models have been applied to model SARS-CoV-2 infections, and the SEIR model is particularly population, since it incorporates an infection incubation period. Recent study has proposed [16] further extensions incorporating separate compartments for asymptomatic and pre-symptomatic cases.

In this paper, we further extend the SEIR model to incorporate testing and errors in testing as visualized in Figure 1 The novelty of this paper is in the direct incorporation of the high false negative rates of COVID-19 RT-PCR tests into the model structure. We break the infectious compartment in two, separating out untested and tested infectious cases. It will be natural to assume that the a large proportion of the untested node consists of the asymptomatic individuals. But it also contains a proportion of symptomatic people also. But the majority of the untested node is the asymptomatic people. For tested individuals, we consider separate compartments for positive (“Tested Positive”) and negative (“False Negative”–truly infected by falsely reported as negative) test results. We assume that only people that have been exposed will test positive, so the Tested Positive compartment corresponds to true positives. Our model specification excludes a small fraction of people who are diagnosed based on only symptoms, without ever having a diagnostic test. After disease exposure and possibly testing, infectious people will then either die or recover from disease. These two outcomes are separated out into compartments corresponding to reported and unreported cases. We include both untested individuals and individuals with false negative tests in the unreported compartments.

**Figure 1:**
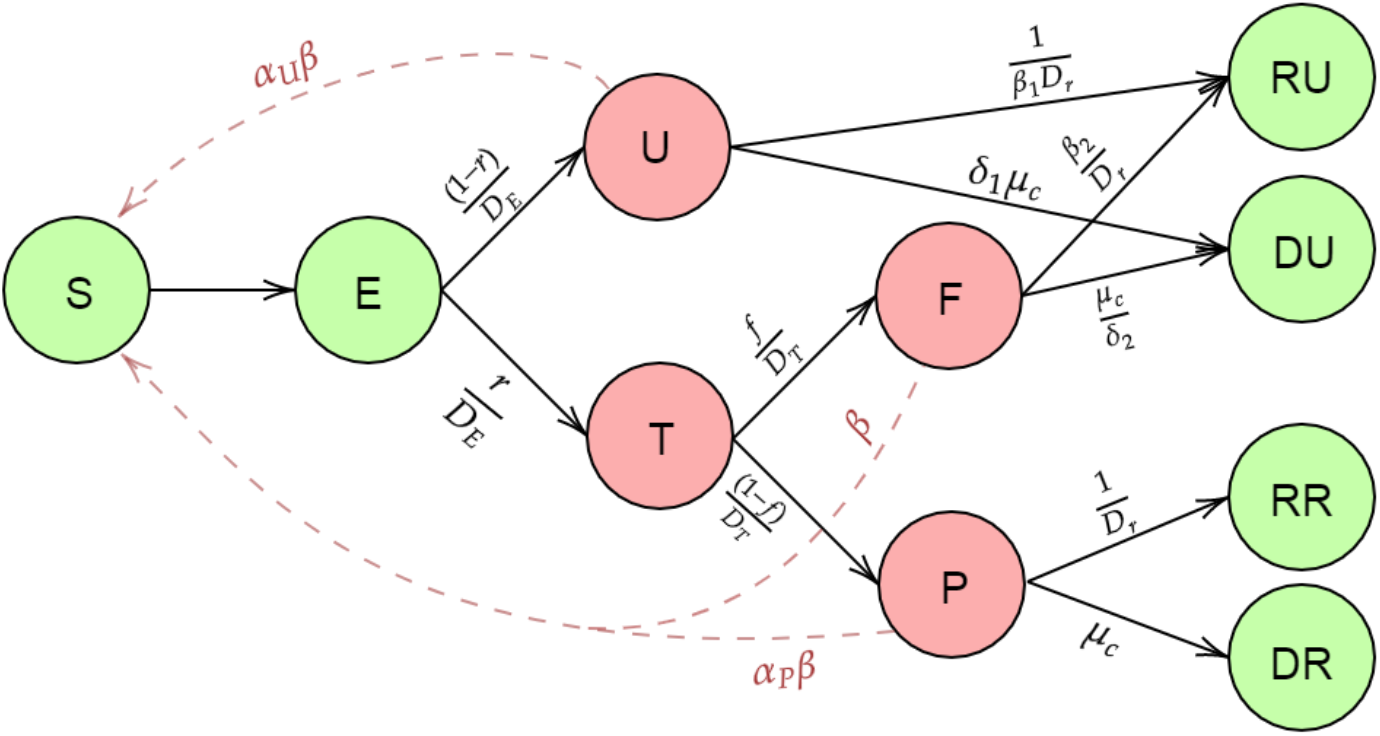
Compartmental model incorporating false negative test results

We divide the entire population into 10 main compartments as described in Figure 1: *S* (Susceptible), *E* (Exposed), *T* (Tested), *U* (Untested), *P* (Tested positive), *F* (Tested False Negative), *RR* (Reported Recovered), *RU* (Unreported Recovered), *DR* (Reported Deaths) and *DU* (Unreported Deaths).

To avoid any confusion we briefly describe our compartments as follows:

- **Susceptible (***S*): Individuals not yet exposed to the disease.
- **Exposed (***E*): Individuals who have been exposed to the virus but have not started spreading infection.
- **Untested (***U***) :** After a certain time (incubation period), infected individuals start spreading the disease. Infectious individuals who are never tested after their incubation period are called Untested.
- **Tested (***T*): Infectious individuals who undergo laboratory testing.
- **Positive (***P*): Tested infected individuals who are reported as COVID positive, called positives.
- **False negative (***F* ): Tested infected individuals who are not reported, called false negatives.
- **Recovered unreported (***RU* ): Unreported patients who recovered from COVID-19.
- **Recovered reported (***RR*): Reported patients who recovered from COVID-19.
- **Deceased unreported (***DU* ): Deaths due to COVID-19 among unreported cases.
- **Deceased reported (***DR*): Deaths due to COVID-19 among reported cases.

### 2.1 Compartmental Parameters

We assume that the time a person stays in a particular compartment/node follows an exponential distribution with a rate. In theory, transitions can happen in continuous time, but for practical implementation we discretize time into days. Below, we define the main parameters of our model (shown in Figure 1):

- *β* : Rate of transmission of infection by false negative individuals.
- *α*_*p*_ : Ratio of rate of transmission by tested positive patients relative to false negatives. We have assumed
- *β*_*p*_ < 1, since patients who are tested positive are likely to adopt isolation measures, where the chance of spreading the disease is less than that of false negative patients who are mostly unaware of their infectious status.
- *β*_*u*_ : Scaling factor for the rate of transmission by untested individuals. *β*_*u*_ is assumed to be < 1 as compartment U mostly consists of asymptomatic or mildly symptomatic cases who are on average likely to be less contagious than those having symptoms.
- *D*_*e*_ : Incubation period (in number of days).
- *D*_*r*_ : Mean number of days till recovery for those who test positive.
- *D*_*t*_ : Mean number of days for the return of test result.
- *μ*_*c*_ : Death rate due to COVID-19 infection which is equivalent to the inverse of the average number of days from disease onset to death times the true infection fatality rate.
- *λ, μ* : Natural birth and death rates in the population. These are assumed to be equal for the sake of simplicity.
- *r* : Probability of being tested for infection, akin to the ascertainment rate used in other comparable SEIR-type models.
- *f* : False negative probability of RT-PCR test.
- *β*_1_ and 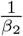 : Scaling factors for rate of recovery for undetected and false negative individuals respectively. Both *β*_1_ and *β*_2_ are assumed to be less than 1. The severity of symptoms in untested individuals is assumed to be less than those tested positive. Consequently, untested individuals are assumed to recover faster than those who tested positive. The time to recovery for false negatives is assumed to be larger than those who tested positive since their absence of diagnosis and consequently formal hospital treatment.
- *δ*_1_ and 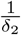 : Scaling factors for death rate for untested and false negative individuals respectively. Both *δ*_1_ and *δ*_2_ are assumed to be less than 1. The untested individuals are assumed to have a smaller probability of dying relative to those who test positive, since untested people are mostly asymptomatic. False negatives are assumed to have a higher probability of dying relative to those who test positive due to absence of diagnosis and consequently formal hospital treatment.

We briefly describe the transmission dynamics of our model. First, the Susceptible (S) people are coming in contact with one infected individual at a given time at the four infectious compartments/nodes U, T, F and P with rates *α*_*u*_*β, α*_*p*_*β, β* and *β* respectively. After getting infected they move to the Exposed Node. After the incubation period, they move into the Untested (U) and the Tested (T) node with rates 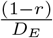 and 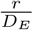. Those who are tested are reported to be positive or negative after *D*_*t*_ days with rate 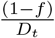 and 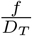. Those in the Untested compartment move to the the Recovered Unreported Node (RU) and the Death Unreported Node (DU) with rates 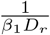 and *δ*_1_*μ*_*c*_ while the Tested positive people move to the Recovered Reported Node and Death Reported Node with rates 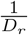 and *μ*_*c*_ respectively. Finally, the Tested False Negative people (F) move to the the Recovered Unreported (RU) and Death Unreported (DU) with rates 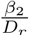 and 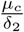 respectively. Let *S*(*t*), *E*(*t*), *T*(*t*), *U*(*t*), *F*(*t*), *RR*(*t*), *RU*(*t*), *DR*(*t*) and *DU*(*t*) denote the number of people in each the compartments described above at the time-point *t*.

### 2.2 Differential Equations

The number of individuals at time-point t at each node follows the set of differential equations described below. To simplify our model, we assume that there is no delay in receiving test results once a person becomes infectious (so *D*_*t*_ = 0). The differential equations corresponding to non-instantaneous testing have been given in the section 1 of Supplementary materials. The following are the differential equations corresponding to instantaneous testing which we have used for our analysis.

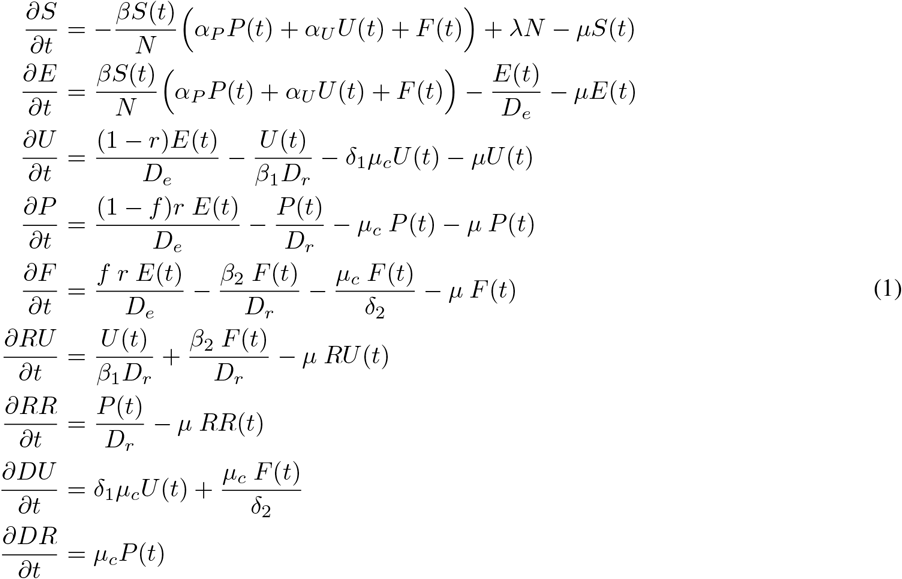

### 2.3 Basic reproduction number

The basic reproduction number (or reproductive ratio) is defined as the number of infections that are expected to occur on average in a homogeneous population as a result of infection by a *single infectious individual* when the entire population is susceptible at the start of the pandemic. We calculate the basic reproduction number for the above model using the Next Generation Matrix Method shown in [26]. In the next generation matrix method, we start by calculating the next generation matrix and the spectral radius of the next generation matrix gives us the basic reproduction number. The expression for *R*_0_ comes as

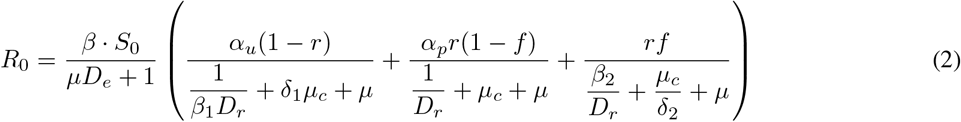

where 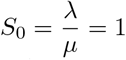 since we have assumed natural birth and death rate to be equal within this short period of time. The detailed derivation is given in the Section 2 of the Supplementary Materials.

#### Special Cases

We develop an intuitive understanding of the above expression by studying some special cases as follows:

- **SIR model :** In the SIR model, there are only 3 compartments : *S* (Susceptible), *I* (Infectious) and *R* (Removed). To obtain the *R*_0_ for SIR model using the expression in (2), we assume *r* = 1, *f* = 0, birth rate (*λ*) = natural death rate (*μ*) = 0 and 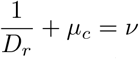 is the removal rate. Here, the death and recovered compartments are merged into one compartment called *R* (Removed). In the SIR model, there is only one infectious component *I* (which is our *P* component), so we assume *α*_*p*_ = 1. With these assumptions, our model reduces to SIR model, and we can simplify the expression in (2) as follows

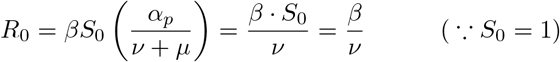

We recover the well-known form of *R*_0_ for the SIR model as derived in [26] as a special case.
- **SEIR model :** In the SEIR model, we have 4 compartments : *S*(Susceptible), *E*(Exposed), *I*(Infectious) and *R*(Removed). To obtain the expression of *R*_0_ for SEIR model, we make all the assumptions as we just made for SIR model except the incubation period *D*_*e*_ = 0. Here we take 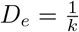. We can also assume non-zero natural birth and death rates *λ* and *μ*. Under these assumptions, the expression of *R*_0_ in (2) becomes

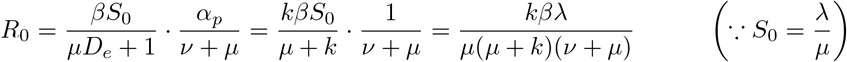

This is the expression of *R*_0_ for the SEIR model as derived in [26] as a special case.

### 2.4 Estimation

Typically, one can solve the system of equations by assuming initialization constraints/values, then fixing certain key parameters and allowing parameters of interest to be estimated based on data. We assume there are two key time varying parameters *β* and *r* in this model capturing transmission rate and ascertainment rate over time. This assumption reflects the natural progression of the pandemic coupled with changes in prevention/testing strategies in the population under study. The steps for estimating the parameters are outlined as follows.

### 2.4.1 Solving the system of Differential Equations

The Differential Equations that have been described in the previous section are for continuous time modelling. We have represented the system at discrete time points by approximating the rate of change/derivative in counts corresponding to any general compartment *X* with respect to time *t* given by 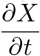 by the difference between that compartment counts on the (*t* + 1)^*th*^ day and the *t*^*th*^ day, i.e. (*X*(*t* + 1) − *X*(*t*)). Starting with initial counts at each node on Day 1 and using the discrete time recurrence relations, we can find the counts for each of the compartments at time *t*. Some of these discrete time recurrence relations are shown below:

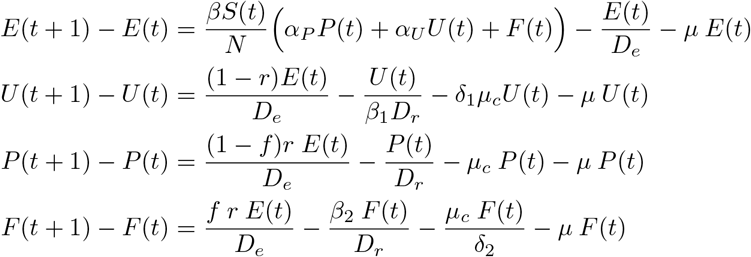

#### 2.4.2 Distributional Assumptions and Likelihood

We assume that the joint distribution of the counts transitioning to each compartment at a given time follows a Multinomial distribution. For example, from the Exposed node, one can move to the Positive, False Negative, or Untested nodes, or they may die due to natural causes. Let *δ*_*X*→*Y*_ denote the number of individuals moving from *X* to *Y* compartment at time *t* with *δ*_*X*→0_ denoting the number of individuals in compartment *X* dying at time *t*. Thus, for the exposed node we have the following stochastic distribution:

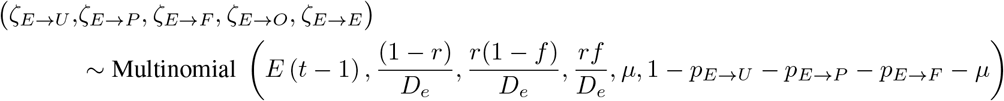

The complete model with the distributions of latent nodes have been described in details in section 5 of Supplementary materials.

We now write down the likelihood that underlies any estimation procedure.

##### Case I: Only using data on daily new cases

For this situation, we assume that given the parameters, the number of new confirmed cases on the the *t*^*th*^ day depends only on the number of exposed individuals on the previous day. Let the number of newly reported cases on day *t, P*_new_(*t*), say, follow a distribution with probability mass function (pmf) *h*(*x* | *β*, **r**, E(*t* 1)). Then, we can write the likelihood of *β* = *β*_1_, *β*_2_, …, *β*_*s*_ and **r** = r_1_, r_2_, …, r_*s*_ where *s* denotes the number of disjoint time periods, as :

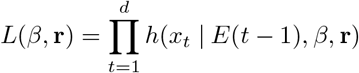

Here, *d* denotes the last day used in model-fitting. We assume that *P*_new_(*t*) follows a Binomial distribution with size *E*(*t* −1) and probability 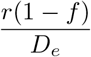. This is a natural corollary of the assumption that counts corresponding to all the compartments jointly follow a Multinomial distribution. Thus the daily number of positive cases marginally will follow a binomial distribution. Alternatively one can assume that *P*_new_(*t*) follows a Poisson distribution with rate 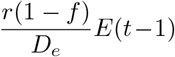, where *E*(*t*−1) is the conditional expectation of the number of exposed at day (*t* −1).

##### Case II: When daily data on new cases, recoveries and deaths are available

Marginally, the distribution for the daily number of positive cases remains the same as before. For the Recovered and Death nodes, the joint distribution is again a multinomial distribution given *P* (t – 1). If *P*_new_, *R*_new_, and *D*_*new*_ follow the distribution with pmf *h* (*x, y, z* | *β*, **r**, E(*t*−1). Then, we can write the likelihood of *β* = {*β*_1_, *β*_2_, …, *β*_*s*_} and **r** = {*r*_1_, *r*_2_, …, *r*_*s*_} where *s* denotes the number of time periods as follows:

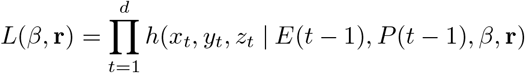

This expression follows from the relationship

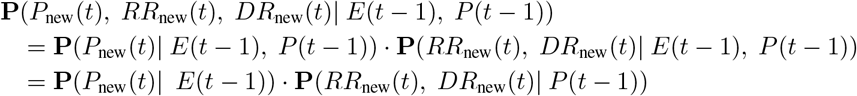

The simplifications in the above equation are consequences of model assumptions. We have

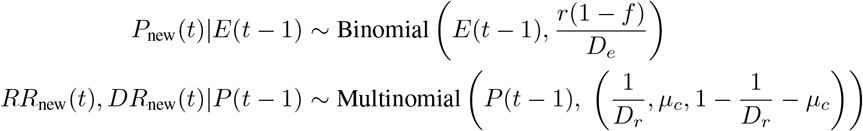

The values of *E*(*t*−1) and *P*(*t*−1) are obtained from solving the discrete time differential equations that have been described in Section 2.4.1. Prediction of the number of active cases is critical from a public health perspective. However, in many countries data on Recoveries and Deaths is not very reliable. In that case, it is better to go with the simpler Poisson or Binomial likelihoods. Following are the respective counts that we are interested in predicting.

Table 1 shows the variables of interest and how they can be obtained from our model. It is important to note that we are modelling the number of active cases, recoveries and deaths first and using them we obtain estimates of cumulative cases. So, predicting active cases, recoveries and deaths accurately is sufficient to obtain good estimates of cumulative cases. Further, the parameters which we estimate (*β* and *r*) determine the number of active cases which in turn determine the number of deaths and recoveries. So, the dependence of deaths and recoveries on the parameters which we estimates is through the number of active cases. Hence, we will judge our models primarily based on their performance in predicting active cases.

**Table 1:** Variables of interest and their expressions in terms of model compartments

| Counts of Interest | Notation (at time t) |
| --- | --- |
| Reported Active Cases | P(t) |
| Unreported Active Cases | U(t)+F(t) |
| Total Active Cases | P(t)+U(t)+F(t) |
| Reported Cumulative Cases | P(t)+RR(t)+DR(t) |
| Unreported Cumulative Cases | U(t)+F(t)+RU(t)+DU(t) |
| Total Cumulative Cases | P(t)+RR(t)+DR(t)+ U(t)+F(t)+RU(t)+DU(t) |
| Reported Deaths | DR(t) |
| Unreported Deaths | DU(t) |
| Total Deaths | DR(t)+DU(t) |

#### 2.4.3 Choice of Priors

We use a Bayesian estimation technique supported by Markov chain Monte Carlo (MCMC) sampling for estimating *β* and **r**. For the parameter *r*, we have assumed a *U*(0, 1) prior distribution while for *β*, we have assumed an improper non-informative flat prior given by :

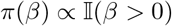

Instead of a non-informative prior, we could have taken a more informative prior. We could alternatively assume a lognormal prior for *β* and *R*_0_. This will induce an implicit prior distribution on r by virtue of the relationship between *R*_0_, *β* and *r*. We opt to induce a non-informative prior on r because we do not have enough information on r, the ascertainment rate.

#### 2.4.4 Posterior Sampling Using MCMC

After specifying the likelihood and the prior distribution, we sample from the posterior distribution of the parameters using an iterative Markov Chain Monte Carlo (MCMC) algorithm. Within each iteration, parameters are drawn using the Metropolis-Hastings method with a Gaussian random walk proposal distribution. We run the algorithm for 100,000 iterations with a burn-in period of 100,000 using `Rstudio` [18]. To reduce autocorrelation of the sampled observations, we use thinning bins of size 100. Finally, the mean of the posterior draws is used as a Bayes estimate of *β* and *r* for the different time periods. For every posterior sample we get am estimated value of *β* and *r*. The counts corresponding to the different compartments at each time point t are obtained by draws from their sampling distribution conditional on the sampled values of *β* and *r*. We repeat this for all *t* in our interval. To obtain a 95% Bayesian credible interval for all parameters we use the 2.5% quantile and 97.5% quantile of the posterior distribution.

## 3 Extensions

With the above structure as our primary analytic foundation, we extend this estimation approach in three primary directions to make the method adapt to real data better based on what we are observing during this pandemic.

### 3.1 Extension 1. Time varying Case-Fatality Rate (mCFR)

After exploring this base model across countries, we observe that the death rates and in turn the case-fatality rates are also changing during the course of this pandemic between and within countries. The usual case fatality rate (CFR) is defined as:

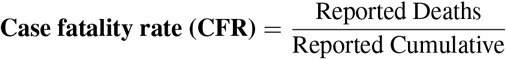

The modified CFR or mCFR includes only the removed cases (deaths+recoveries) in the denominator as the outcomes are known only for this subset of individuals.

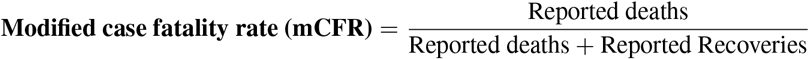

Figure S1 of the Supplementary Section shows that while countries like Belgium, USA, Italy, Spain have very high mCFR, India and Russia have comparatively much lower mCFR. We also note that initially most countries experience a high mCFR and it gradually settles to a lower value as the case counts and recoveries rise. Hence, we hypothesize that modeling mCFR as a time varying quantity will improve the prediction of active cases and deaths.

Thus we introduce a third time varying parameter called the mCFR along with *β* and *r* in a three parameter multinomial model. With this change, the differential equations in (1) will remain the same. We use mCFR as opposed to CFR because in our model we use it to determine what is the probability that an infected person from node *P* moves to the death node (*DR*). The remaining will go to the recovered node *RR*. The new recovery rate will be 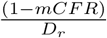, while the new death rate will be mCFR×*μ*_*c*_.

### 3.2 Extension 2. Testing of infectious people based on symptoms

The problem with the base model is that we have implicitly assumed that the probability of a person being tested is equal for all infected individuals. However, that is not the case in reality. The probability of being tested for a truly infected person largely depends on symptoms. On an average, a person with severe symptoms will have the highest probability of being tested followed by the mildly symptomatics and the asymptomatics. We extend the previous misclassification model accounting for symptom dependent testing. We split the **Exposed(E)** compartment into three nodes **Severe Symptomatic (Se), Mild Symptomatic (Mi)** and **Asymptomatic (As)**.

We will here assume that the individuals with severe symptoms will be tested with probability 1 while the mild and asymptomatic ones will be tested with probability *t*_1_ and *t*_2_ respectively. We will also assume the probability of an infected person having severe, mild or no symptoms are *p*_1_, *p*_2_, *p*_3_ respectively. Figure S2 of the Supplementary Section presents the different compartments and their corresponding dynamics for this model.

Now, all the differential equations remain the same except the ones corresponding to the nodes P, U and F. The new set of differential equations corresponding to these three nodes are:

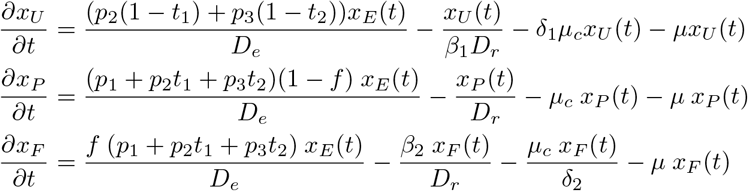

Due to identifiability issues, all the parameters described above cannot be estimated. We assume known values for *p*_1_, *p*_2_ and *p*_3_ that can be obtained from existing data. We also assume that *t*_1_ = *kt*_2_ where k is greater than 1 and known. This assumption implies that the probability of receiving a test for a person with mild symptoms is more than a person with no symptoms. We run a sensitivity analysis for different values of k.

This model is more or less equivalent to the multinomial two parameter model. The only additional information that we are obtaining here is the allocation of tests conditional on symptoms. We essentially have expressed the probability of getting tested or r as the sum of three different probabilities by using the theorem of total probability. Namely,

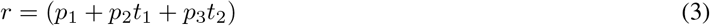

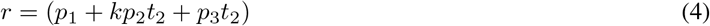

Our main parameters of interest that we are going to estimate now is *β* and *t*_2_ instead of *β* and *r*. Since this model is a simple reparameterization of our original model we do not discuss this any further.

### 3.3 Extension 3. Selection model: Who is getting tested?

So far we have been concerned with only the testing of truly infected individuals. However, symptoms may manifest in an infected individual or an uninfected individual. The cause of symptoms (both mild and severe) in susceptible individuals may be due to respiratory diseases such as influenza and the common cold.

It may be reasonable to assume that each individual, regardless of their underlying true disease status, has a probability of being tested that depends on the symptoms they have (or don’t have). This probability could also depend of other covariates such as job types or pre-existing co-morbidities. For simplicity we will only consider symptoms determining testing for both diseased and disease-free. We want to create an analytic framework to study the selection bias due to testing in the population.

To this end, we consider testing strategies that mandate that individuals with severe symptoms are always tested provided sufficient tests are available. After all the individuals with severe symptoms are tested the remaining tests are divided among those with mild symptoms and asymptomatics according to some given allocation rule that is independent of their true disease status given observed symptoms. We also assume that the number of tests to be performed in a given day does not depend on the true infection counts and is an external input. One advantage of using the number of tests as an input to the model is that we can study how the number of available tests influences the population infection rate in the long term. Figure 2 provides a visualization of this expanded model.

**Figure 2:**
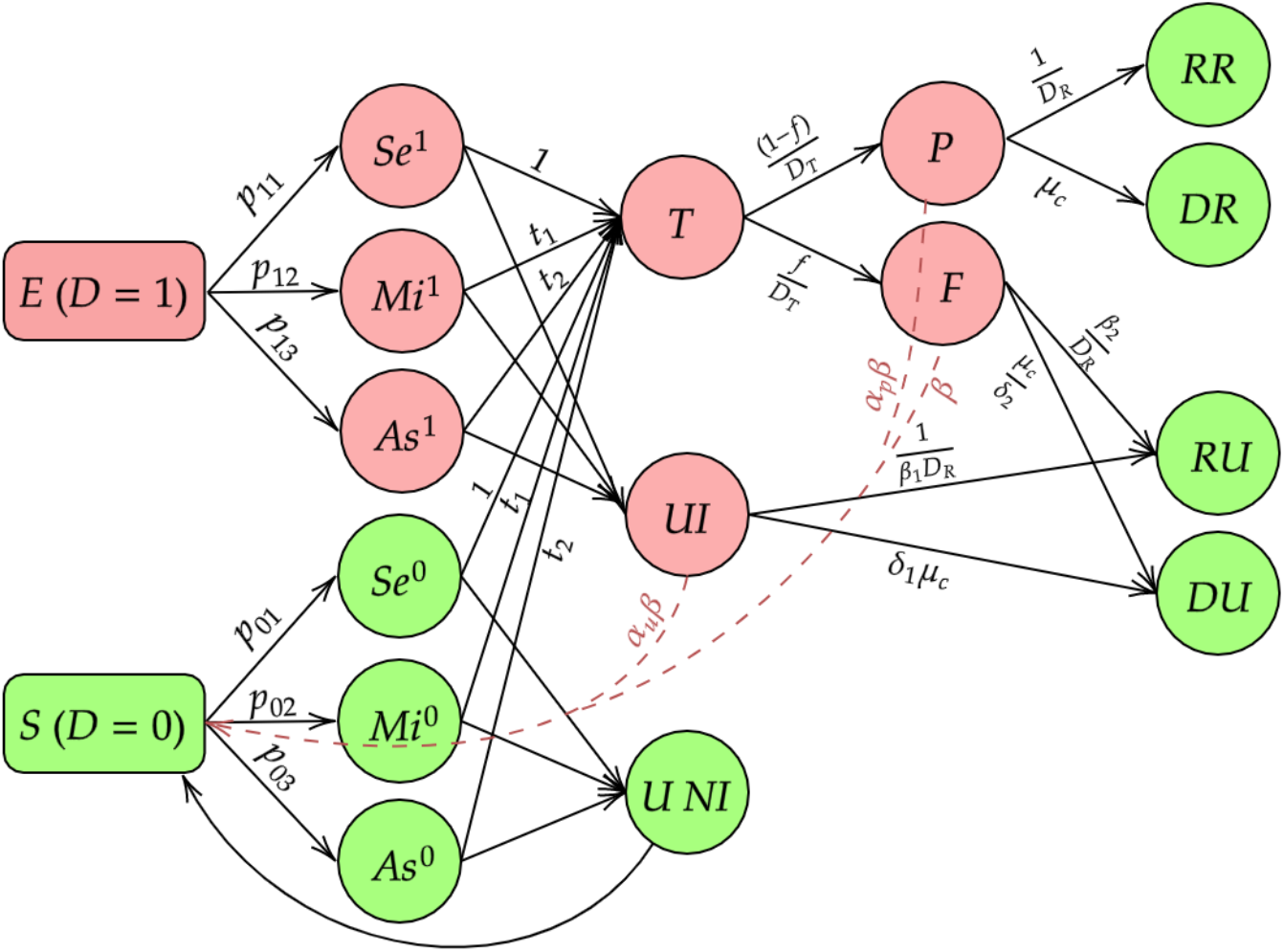
Selection Model

The new compartments are summarized as follows:

- *Se*^1^ : Those who have developed severe symptoms and have a true COVID-19 infection.
- *Mi*^1^ : Those who have developed mild symptoms and have a true COVID-19 infection.
- *As*^1^ : Those who have developed no symptoms and have a true COVID-19 infection.
- *Se*^0^ : Those who have developed severe symptoms and do not have a true COVID-19 infection.
- *Mi*^0^ : Those who have developed mild symptoms and do not have a true COVID-19 infection.
- *As*^0^ : Those who have no symptoms and no COVID-19 infection.
- *UI* : Those who are untested with an active COVID-19 infection
- *UNI* : Those who are untested without an active COVID-19 infection.

The differential equations corresponding to this model have been provided in section 6 of Supplementary materials. Though conceptually appealing and pragmatic, this model has identifiability issues, and estimation requires substantial additional information. In particular, we need to know the mechanism by which people are tested including the corresponding probabilities of testing. Additionally, we need to know the true symptom distributions for the exposed and susceptible people. It will be often quite hard to obtain this information as part of regularly released data sources by countries across the world. Thus, implementation of this model may be wrinkled with too many subjective choices. However, we still think this is a valuable formulation as it helps us to understand, analytically and intuitively, how selection bias can influence our estimates of interest. For example we derive an expression for *R*_0_ under this complex structure. **Basic Reproduction number :** We calculate *R*_0_ using the same Next Generation Matrix Method as in Basic reproduction number. For this new framework that includes uninfected individuals as well as false negatives, we have derived an expression for the basic reproduction number as follows :

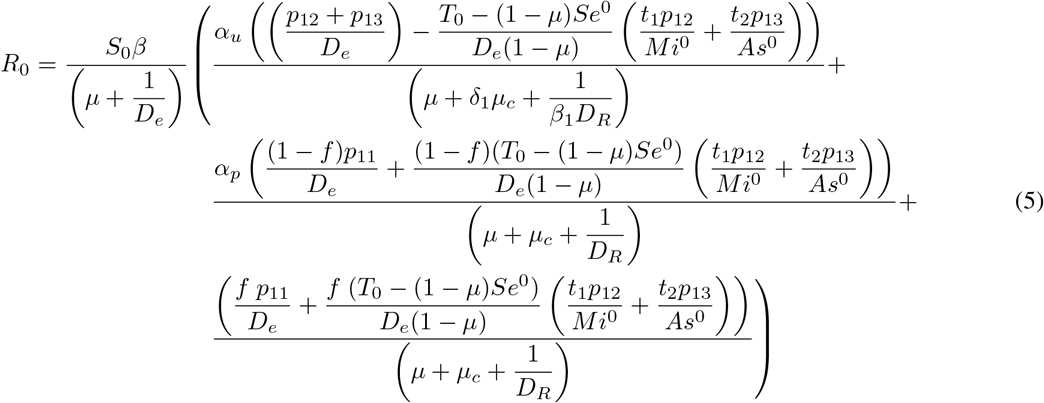

#### Decoupling the effect of selection and misclassification on *R*_0_

To provide some intuition about the effect of selection bias and misclassification on the value of *R*_0_, we consider an example setting where we first compute the value of *R*_0_ from equation (5) with *f* = 0 (no misclassification, selection), *f* = 0.3 (misclassification+selection) using the same set of parameters. To isolate the effect of selection, we consider a model where selection is random (for further details refer to section 8 of supplementary materials) and evaluate *R*_0_ when *f* =0 (no misclassification or selection) and *f* = 0.3 (only misclassification, no selection). We consider a hypothetical population of 1 million people. We set *β* = 0.25, *r* = 0.1, *p*_0_ = (10^−6^, 10^−5^, 1 − 10^−6^ − 10^−5^) *p*_1_ = (0.02, 0.18, 0.8), *t*_1_ = 0.7 and *t*_2_ = 0.3. We consider three different values of the number of tests per thousand population (0.1, 0.5, 1.0, 2.0). These values are consistent with what we have seen across the world. For example, as on June 30, India and US are doing 0.15, 1.82 daily tests per thousand population. The rest of the parameters are same as in (4.1). The following table (2) shows the values of *R*_0_ for the four configurations. From table (2), we conclude that under random selection, *R*_0_ is not sensitive to the total number of tests and false negatives and remains around 1.54, the true value. The values in the 3^rd^ column are inflated, which indicates a substantial effect of selection bias on *R*_0_, especially when the number of tests are large, even when the tests are perfect. The fourth column underpins the key issue that the *R*_0_ can be very far from the true value with both selection bias and misclassification, especially with large number of tests being distributed in a non-uniform way.

**Table 2:** Effect of misclassification and selection bias on *R*_0_

| Number of tests<br>per thousand | Model |  |  |  |
| --- | --- | --- | --- | --- |
|  | No Selection Bias |  | Selection Bias |  |
| | $f = 0$ | $f = 0.3$ | $f = 0$ | $f = 0.3$ |
| 0.1 | 1.54 | 1.53 | 1.64 | 2.09 |
| 0.5 | 1.54 | 1.54 | 2.02 | 4.08 |
| 1.0 | 1.54 | 1.54 | 2.50 | 6.56 |
| 2.0 | 1.54 | 1.55 | 3.45 | 11.54 |

## 4 Analysis of COVID-19 Pandemic in India

Now that we have described our models and methods, we proceed to assess their performance with real and simulated data. For this purpose, we fit our models to daily new reported case, recovery and death-counts in India from 1^st^ April to 30^th^ June. We have chosen India because the pandemic has been prevailing in India for a very long time and the incidence curve has not turned corner as of August 31. With more than 3.5 million total reported cases, approximately 75,000 new cases and 1000 deaths reported per day at the end of August, India presents a unique setting to assess our models. Another important feature for India is that the national lockdown periods are clearly defined, which helps us to define the time changes in *β*. For most of the analysis, we have used our Multinomial 2 parameter model with only misclassification, since symptom-dependent testing data are not available for India. We have used the data for the whole country as well as for the states with some of the highest incidence of the disease namely Delhi and Maharashtra to characterize the heterogeneity. The data are available at a crowd-sourced dashboard covid19india.org. We compare our models’ prediction with reported counts for total cumulative, reported deaths and reported active cases in India. The unreported compartments cannot be validated with real data. We split the data into train and test sets with 1^st^ April to 30^th^ June serving as our training period and 1^st^ July to 31^th^ August serving as our test period. The period is divided into time intervals based on public health interventions rolled out in India (see Table 3).

**Table 3:** Phases of public health interventions in India

| Phase | From | To | Salient features |
| --- | --- | --- | --- |
| **Lockdown 1** | 1<sup>st</sup> April | 14<sup>th</sup> April | Complete lockdown in 82 district across the country where confirmed cases were reported. Inter state movement was closed |
| **Lockdown 2** | 15<sup>th</sup> April | 3<sup>rd</sup> May | Extension of lockdown with relaxations in regions with lower spread from 20<sup>th</sup> April |
| **Lockdown 3** | 4<sup>th</sup> May | 17<sup>th</sup> May | Imposition of strict nationwide lockdown |
| **Lockdown 4** | 18<sup>th</sup> May | 31<sup>st</sup> May | NMDA extends lockdown till end of May in all states |
| **Unlock 1.0** | 1<sup>st</sup> June | 30<sup>th</sup> June | Lockdown lifted from many regions while strict lockdowns imposed in containment zones. Services were resumed from 8<sup>th</sup> June. |
| **Unlock 2.0** | 1<sup>st</sup> July | 31<sup>st</sup> July | Lockdown measures were only imposed in containment zones. In all other areas, most activities were permitted. |
| **Unlock 3.0** | 1<sup>st</sup> August | 31<sup>st</sup> August | Unlock 3.0 for August 2020 removed night curfews from 5<sup>th</sup> August. Educational institutions will remain closed till 31<sup>st</sup> August. |

### 4.1 Initial values and parameter setting

We use observed counts on April 1 as *P*(0), *RR*(0) and *DR*(0), the initial values for the reported compartments while the counts in the unobserved compartments are set proportionately to the observed ones. Namely, we have assumed that *E*_0_ = 3(*U*_0_ + *P*_0_ + *F*_0_). The false negative rate is set at *f* = 0.3 and initial value of the ascertainment rate *r* is set at 0.15. The value of *f* is taken based on the reported false negative rates for RT-PCR tests [14]. We assume all the parameters in our model except r and *β* remain constant through the entire course of the disease. We set the latency period *D*_*e*_ = 5.2 days assuming that the latency period is equal to the incubation period. This value has been taken from the estimates in Wuhan [15]. We assume *D*_*r*_ = 17.8 days following the report by WHO and set this as the average time till death for deceased COVID patients. We set 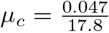 considering the proportion of deaths among the removed persons is about 4.7% in India on June 30. The natural birth and death rates are assumed to be equal. 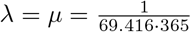 using the fact that average life span of Indians is 69.416 years. The various scaling factors were set as *α*_*p*_ = 0.5, *α*_*u*_ = 0.5, *β*_1_ = 0.6, *β*_2_ = 0.7, *δ*_1_ = 0.3, *δ*_2_ = 0.7. With these values and the time periods described in Table 3, we estimate the *R*_0_ in each period and also predict the counts in the test period using the parameters estimated in the last time period in the training set. We study the effect of these choices for initial values and fixed parameters on our estimable quantities of interest through an extensive sensitivity analysis in the Section 6(Sensitivity Analysis) section.

### 4.2 Basic Reproduction Number

Here Extension 1 and 2 are the Multinomial 3-parameter and Multinomial Symptoms models respectively. We begin with estimates of *R*_0_ by the different models. From Table (4), we note that estimates of *R*_0_ from all of the models are qualitatively similar though there are numerical differences. The estimates of *R*_0_ decreased steadily with time from 3.74 in the 1^st^ period to 1.61 in the 5^th^ period. This shows that the lockdown has been effective in reducing the rate of spread of the disease. The estimated *R*_0_ with accompanying 95% credible intervals from the Multinomial-2-parameter models are shown in Figure 3. The green line indicates *R*_0_ = 2. We observe that from 3^rd^ period onwards the value of *R*_0_ remains below the green line indicating slowing down of the spread of the disease. However it still remains above 1 which means India is yet to achieve herd immunity.

**Table 4:**
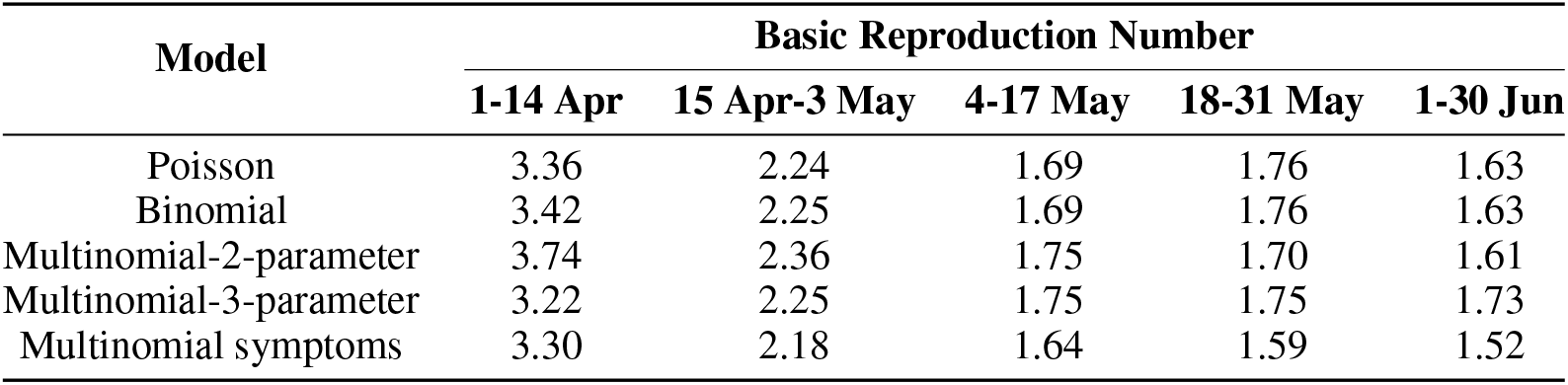
Estimates of *R*_0_ by different models

| Model | Basic Reproduction Number |  |  |  |  |
| --- | --- | --- | --- | --- | --- |
|  | 1-14 Apr | 15 Apr-3 May | 4-17 May | 18-31 May | 1-30 Jun |
| Poisson | 3.36 | 2.24 | 1.69 | 1.76 | 1.63 |
| Binomial | 3.42 | 2.25 | 1.69 | 1.76 | 1.63 |
| Multinomial-2-parameter | 3.74 | 2.36 | 1.75 | 1.70 | 1.61 |
| Multinomial-3-parameter | 3.22 | 2.25 | 1.75 | 1.75 | 1.73 |
| Multinomial symptoms | 3.30 | 2.18 | 1.64 | 1.59 | 1.52 |

**Figure 3:**
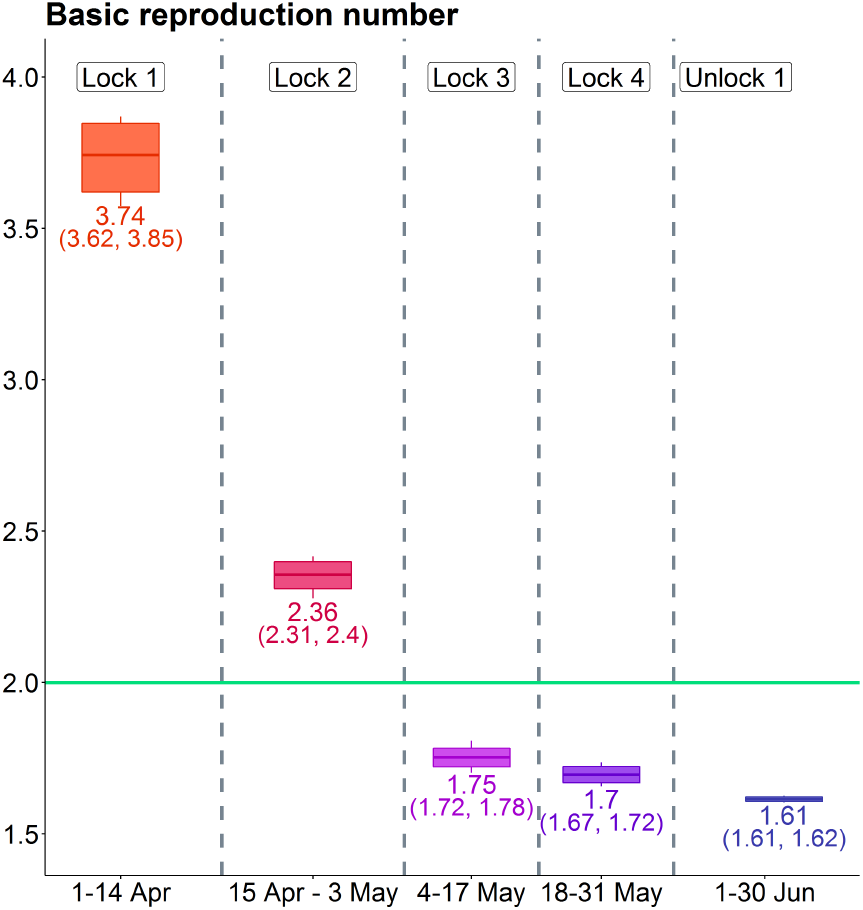
Estimates of *R*_0_ in India across phases. The mean and 95% credible intervals (in parentheses) are provided under the Multinomial-2-parameter model

### 4.3 Prediction accuracy for reported counts

To assess the performance of different models, we compare their predictions of reported active and reported cumulative cases and reported deaths with observed data. We consider the following metric.

#### Mean Squared Relative Prediction Error (MRPE)

The number of reported cumulative start at a number below 5000 on 1^*st*^ April and increase to 3 million in August. As such, we need a measure of error which is scale independent. One such measure is MRPE which is defined as

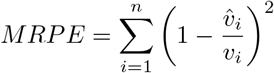

for observed data *v* = (*v*_1_, *v*_2_, …, *v*_*n*_) and predicted vector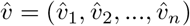.

We present MRPE for each of the models evaluated both on the training and test set. The MRPE is multiplied by a factor of 10, and the lower the value the better.

Table 5 presents the values of MRPE for Total Cumulative, Reported Deaths and Reported Active cases for all the five models. The column-wise minimum are indicated in bold letters for the training set and the minimum in columns corresponding to testing set are highlighted in red. Following is a summary:

**Table 5:** MRPE of different models on training and test set

| Model | Reported Cumulative Cases |  | Reported Deaths |  | Reported Active Cases |  |
| --- | --- | --- | --- | --- | --- | --- |
|  | Train | Test | Train | Test | Train | Test |
| Poisson | **0.066** | 0.098 | 0.948 | 1.235 | **0.357** | 0.749 |
| Binomial | 0.069 | 0.100 | 0.961 | **1.222** | 0.363 | 0.737 |
| Multinomial-2-parameter | 0.206 | 0.087 | 1.248 | 1.270 | 0.569 | **0.705** |
| Multinomial-3-parameter | 0.127 | **0.017** | **0.396** | 1.516 | 0.445 | 1.096 |
| Multinomial symptoms | 0.120 | 0.019 | 1.188 | 1.867 | 0.437 | 1.232 |

- The Poisson and Binomial models perform very similarly. This is expected with large case-counts, the Binomial likelihood approaches the Poisson likelihood.
- We can note that the Multinomial models are doing better in test data in predicting reported cumulative Cases, while in the training data, the Poisson and Binomial models are doing better. The Multinomial 3-parameter and the Multinomial Symptoms models are doing the best in predicting reported cumulative cases in the test data.
- The Multinomial-2-parameter is doing well in predicting reported cumulative cases, reported deaths and reported active cases. The Multinomial-2-parameter model predicts reported active cases better than rest of the models. In the test data, the Multinomial-2-parameter model outperforms the Multinomial-3-parameter model. Though it might seem surprising at first, if we take a deeper look at this matter, we can find a reasonable explanation. If we look at Figure S1 of the Supplementary Section, we can observe that the mCFR for India is almost constant for a large number of days. So we do not need to model mCFR as a time varying parameter as we have done in the Multinomial-3-parameter model. While Multinomial-3-parameter model does well in the training period in case of deaths, the Multinomial-2-parameter model with constant mCFR outperforms the former in the test period. Again, Multinomial-3-parameter model is doing better than the Multinomial-2-parameter in case of Reported Cumulative, but it is doing much worse than the same in case of Reported Active. As reported active is of more importance from the perspective of health care and the Multinomial-2-parameter not only has the least MRPE in predicting reported active cases in the test period, but also does reasonably well in all the test data, we have chosen the Multinomial-2-parameter model for further analysis.

Figure 4 provides the daily prediction trajectories for reported active cases and reported deaths from April 1 to August 31. We focus on reported active cases as the accuracy of this prediction can ably inform health care needs on a daily basis.

**Figure 4:**
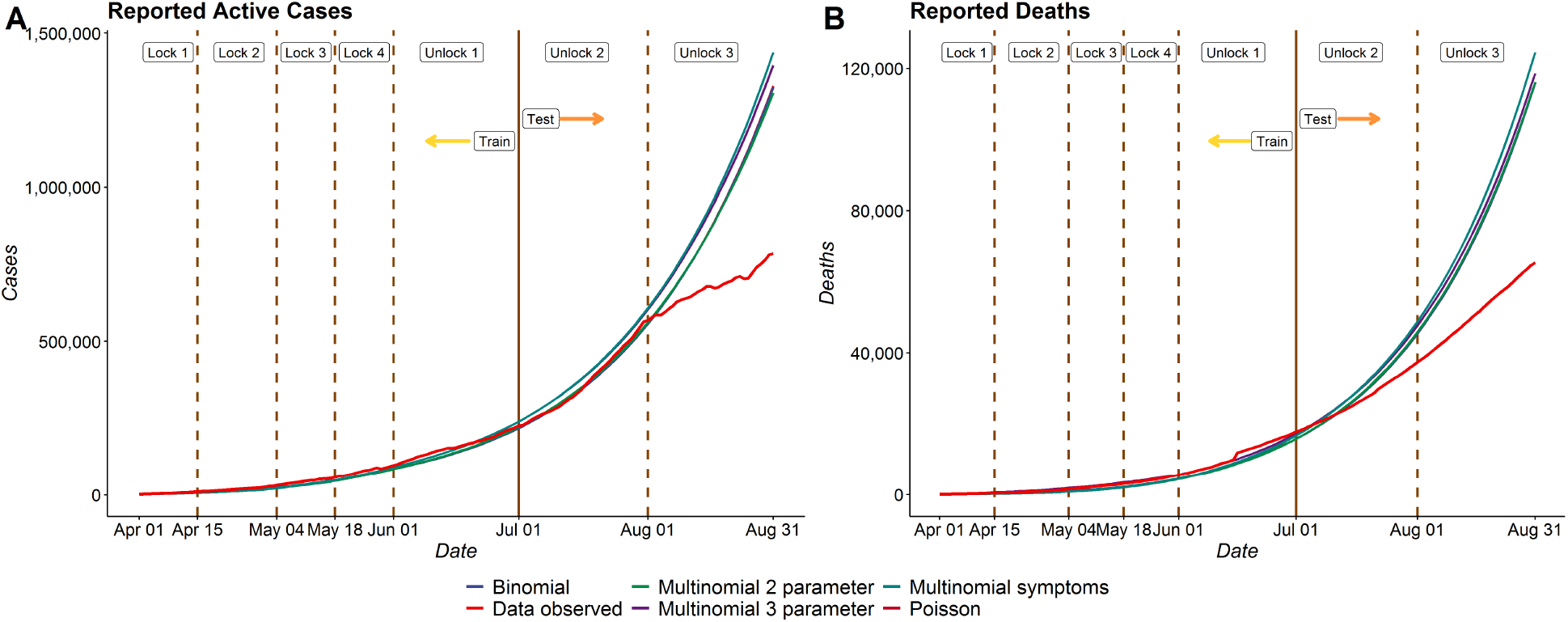
Reported Active Cases in India - Comparison between different models

#### 4.3.1 Prediction of reported and unreported counts

Figure 5 presents the daily composition of active and cumulative COVID cases in India in terms of reported and unreported infections. This figures shows the actual number (left) and proportion (right) of cases who are reported, false negatives or remain untested. We can see from sub-figures (B) and (D) that the proportion of reported active cases and reported cumulative cases with total active and total cumulative cases have increased on an average from April 1 to August 31. The proportion of reported cases among active cases has decreased from 0.12 on 1^st^ April to 0.045 around 15^th^ April. From then, we see it remains fairly constant and increased slightly to 0.048 on 31^st^ August. It is important to note that in spite of enhancing testing and contact tracing, our estimates suggest that about 95% of cases in India remain unreported as of 31^st^ August. In other words, roughly one out of twenty cases is reported. The predicted proportion of reported deaths is roughly 0.17 on August 31, meaning approximately 1 in 6 deaths is reported in India.

**Figure 5:**
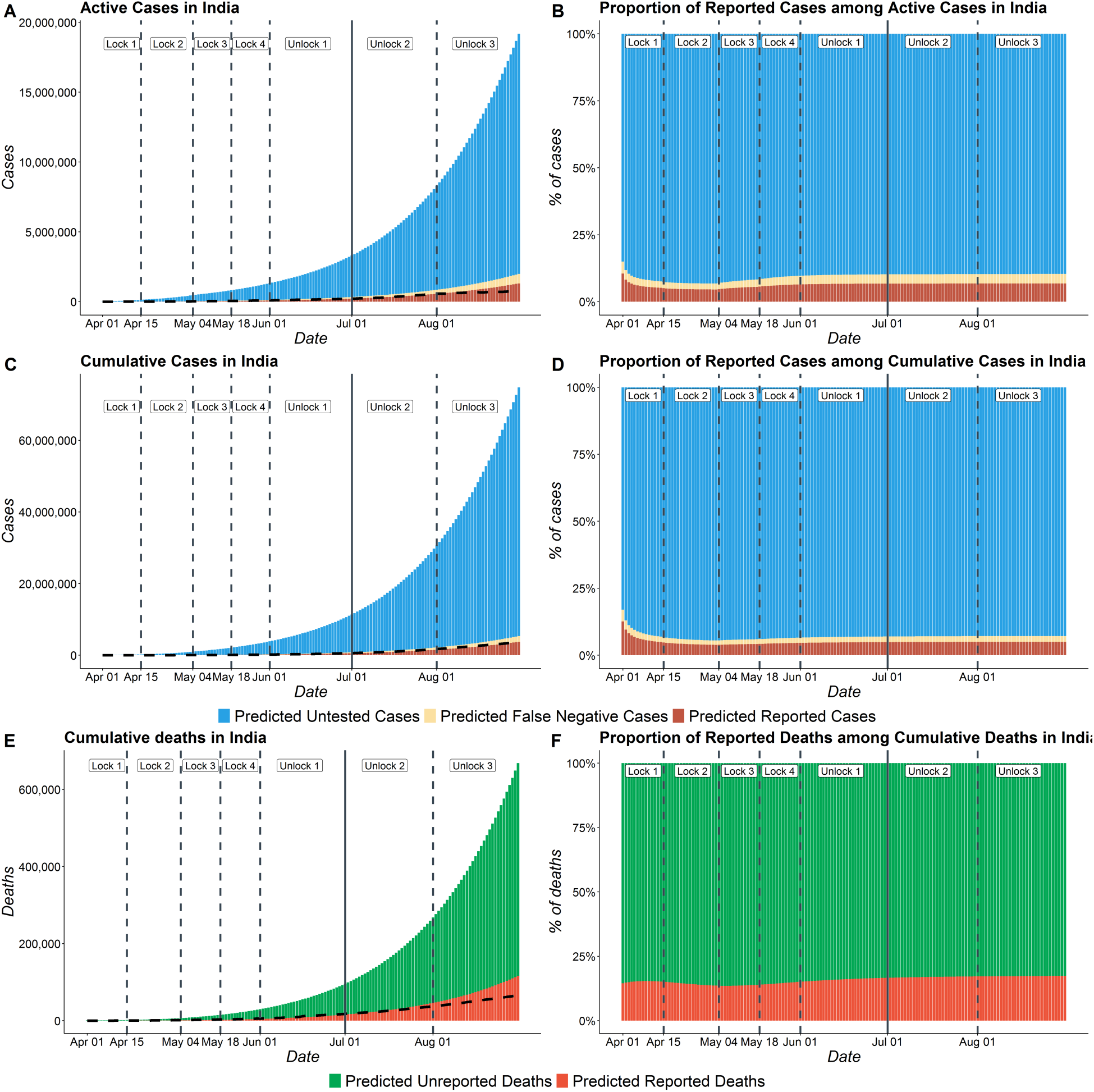
COVID cases in India with number of Reported, False Negative and Untested cases. (A) Total active COVID cases in India from April 1 to August 31 including reported active cases, false negatives active and untested active cases. (B) Proportion of reported active cases among Active COVID cases in India (C) Total cumulative cases in India from April 1 to August 31 including reported cumulative cases, cumulative false negatives and untested cumulative cases. (D) Proportion of reported cases among total cumulative COVID cases in India. (E) Total deaths in India from April 1 to August 31 including reported and unreported deaths. (F) Proportion of reported deaths among total deaths in India. The dotted line in subfigures A, C and E represent the observed data.

#### 4.3.2 Effect of misclassification on prediction

For our main analysis, we assumed the false negative rate of RT-PCR tests to be 30%. Since there is uncertainty in this false negative rate for the different tests rolled out in India, we study how the predictions change for different values of false negative rates (*f* = 0, *f* = 0.15 and *f* = 0.3). From sub-figure (B) in Figure 6, we note that predictions from all the 3 models with different false negative rates coincide with each other for reported active cases as expected. We also note that each of them fit the observed data quite well up to 1^st^ August. Sub-figure (A) of Figure 6 shows that the estimates of total active cases (reported+false negatives+untested) vary substantially across the three assumed values of *f*. As expected, the model with *f* = 0.3 estimates the highest number of unreported cases as it considers the highest false negative rate. The estimates from the model with *f* =0 are about two thirds of that of model with *f* = 0.3. On the other hand, the predictions of model with *f* = 0.15 are only slightly higher than that of model with *f* = 0.

**Figure 6:**
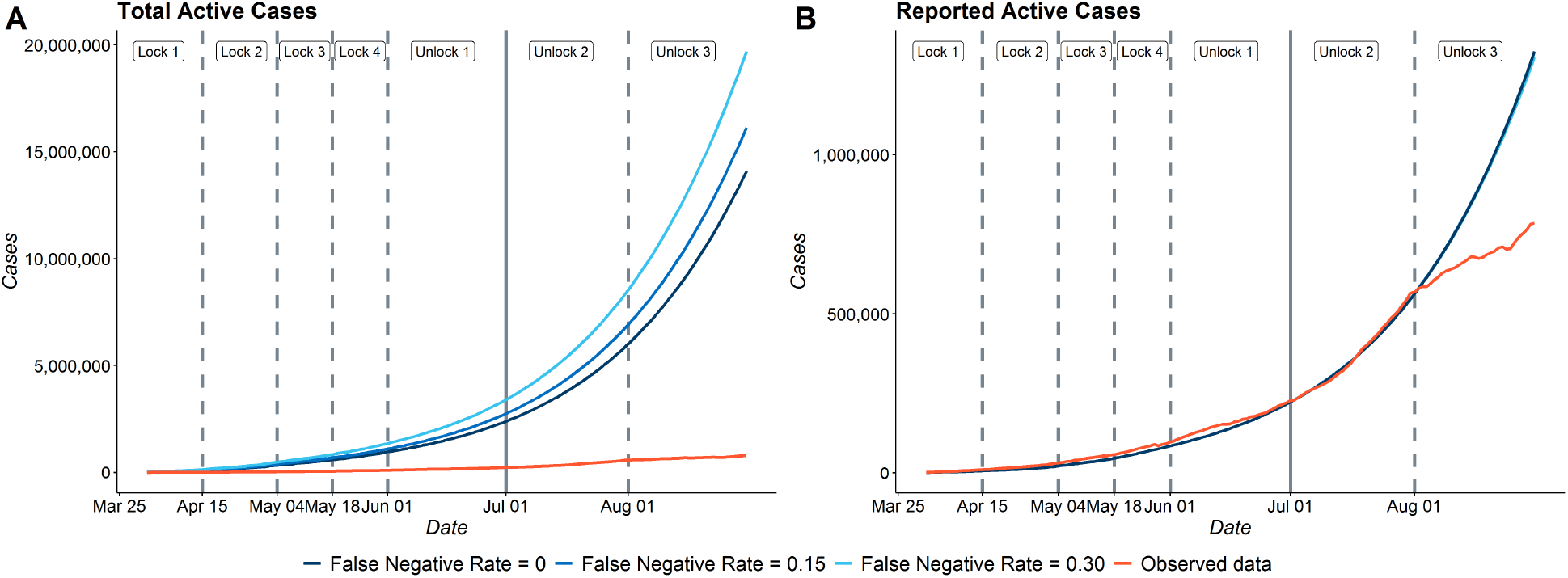
Effect of misclassification on estimates for India (A) Estimates of Total Active Cases for *f* = 0, 0.15 and 0.3 (A) Estimates of Reported Active Cases for *f* = 0, 0.15 and 0.3 with the observed data

### 4.4 Results for Delhi and Maharashtra

There is tremendous heterogeneity in the virus curves across time in India. In fact, the 10 states Maharashtra, Tamil Nadu Delhi, Telengana, Karnataka, Andhra Pradesh, Uttar Pradesh, Gujarat, West Bengal and Bihar constitute 90% of the total cumulative cases of India as of August 31, 2020. We focus on two of the worst-hit states in India - Delhi and Maharashtra.

The state of Maharashtra has nearly 0.8 million reported cumulative cases by 31^st^ August which is more than 20% of total cases in India by that date. On the other hand, New Delhi (national capital of India) had 174,748 cases by 31^st^ August, but the curve turned the corner in June.

We estimate the basic reproduction number for Maharashtra and Delhi and also provide a 62 day prediction of reported active cases. We note that in both the states, the estimates of *R*_0_ have decreased with time. The estimates of *R*_0_ for Delhi have decreased from 5.89 in the 1^st^ period to 1.22 in the 6^th^ period. For Maharashtra, the estimates of *R*_0_ have decreased from 3.58 in the 1^st^ period to 1.51 in the 5^th^ period.

Figure 7 shows the 62-day predictions of reported active cases of Delhi and Maharashtra. Figure 7 shows that our model fits the training data for both the states reasonably well. However, in the test data, we observe that our model overpredicts number of reported active cases for Delhi and Maharashtra. This is due to the sudden decrease in the number of new cases in July and August. Subfigure (A) shows that though our model is overpredicting, it has been able to capture the peak of reported active cases in Delhi in the first week of July.

**Figure 7:**
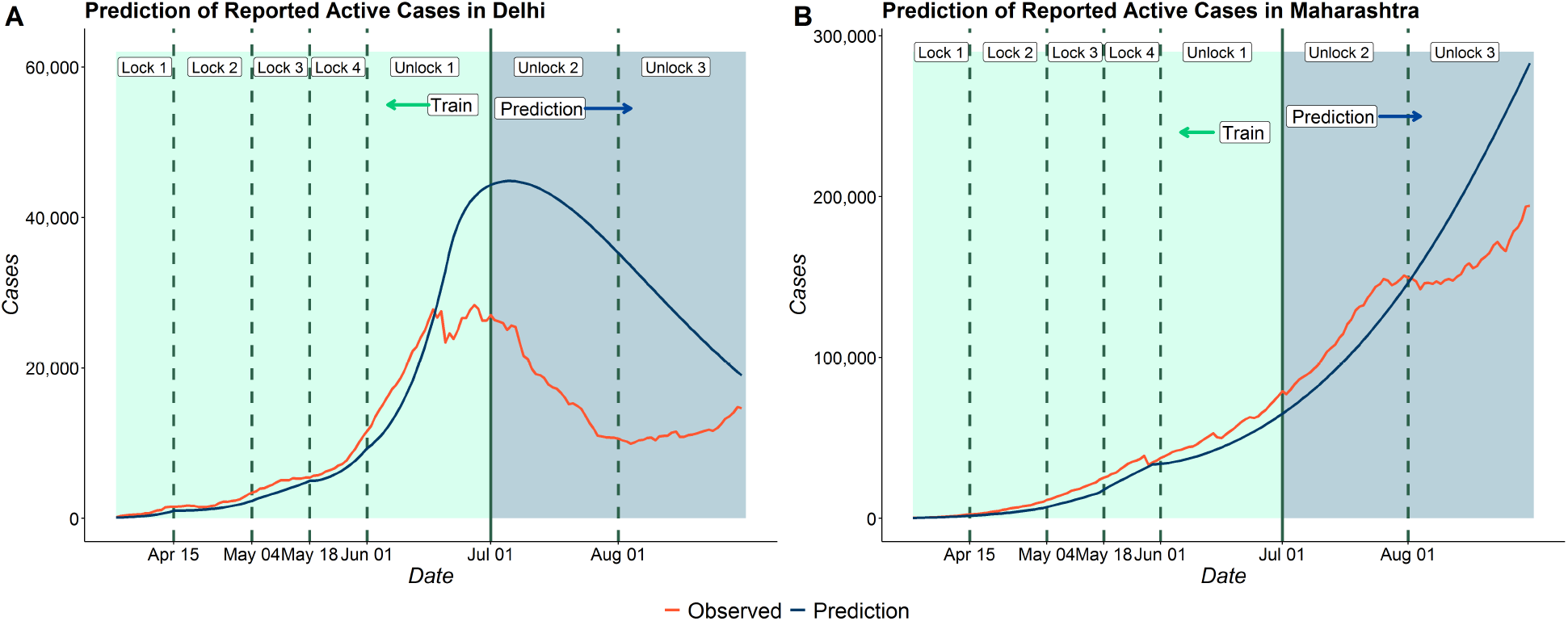
62 day predictions for (A) Delhi and (B) Maharashtra

Table 6 presents a comparison of the under-reporting factors in India, Delhi and Maharashtra for cases and deaths. There is tremendous heterogeneity between states, with case underreporting factors of approximately 20, 53, and 14 and death underreporting factors of 6, 12, and 4 in India, Delha, and Maharashtra respectively. With f =0 these underreporting factors reduce to 12, 35 and 11 for cases and 3, 8 and 3 for deaths in India, Delhi and Maharashtra respectively. This shows that even without accounting for false negatives there possibly exists a large degree of underreporting for case and death counts in India.

**Table 6:**
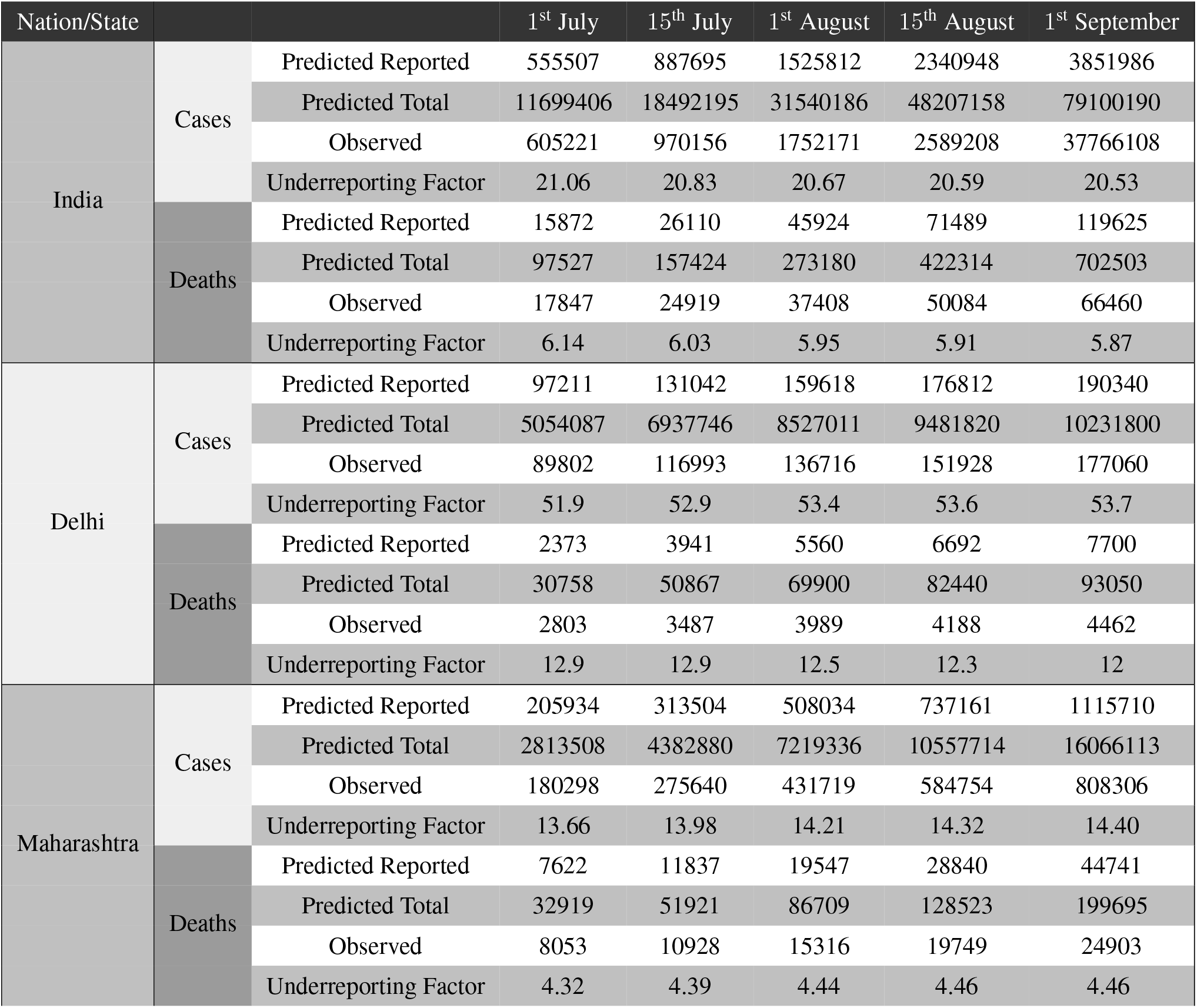
Predicted Cumulative Cases and Deaths (Reported and Total) of India, Delhi and Maharashtra along with observed counts and predicted underreporting factors

| Nation/State |  |  | 1<sup>st</sup> July | 15<sup>th</sup> July | 1<sup>st</sup> August | 15<sup>th</sup> August | 1<sup>st</sup> September |
| --- | --- | --- | --- | --- | --- | --- | --- |
| India | Cases | Predicted Reported | 555507 | 887695 | 1525812 | 2340948 | 3851986 |
|  |  | Predicted Total | 11699406 | 18492195 | 31540186 | 48207158 | 79100190 |
|  |  | Observed | 605221 | 970156 | 1752171 | 2589208 | 37766108 |
|  |  | Underreporting Factor | 21.06 | 20.83 | 20.67 | 20.59 | 20.53 |
|  | Deaths | Predicted Reported | 15872 | 26110 | 45924 | 71489 | 119625 |
|  |  | Predicted Total | 97527 | 157424 | 273180 | 422314 | 702503 |
|  |  | Observed | 17847 | 24919 | 37408 | 50084 | 66460 |
|  |  | Underreporting Factor | 6.14 | 6.03 | 5.95 | 5.91 | 5.87 |
| Delhi | Cases | Predicted Reported | 97211 | 131042 | 159618 | 176812 | 190340 |
|  |  | Predicted Total | 5054087 | 6937746 | 8527011 | 9481820 | 10231800 |
|  |  | Observed | 89802 | 116993 | 136716 | 151928 | 177060 |
|  |  | Underreporting Factor | 51.9 | 52.9 | 53.4 | 53.6 | 53.7 |
|  | Deaths | Predicted Reported | 2373 | 3941 | 5560 | 6692 | 7700 |
|  |  | Predicted Total | 30758 | 50867 | 69900 | 82440 | 93050 |
|  |  | Observed | 2803 | 3487 | 3989 | 4188 | 4462 |
|  |  | Underreporting Factor | 12.9 | 12.9 | 12.5 | 12.3 | 12 |
| Maharashtra | Cases | Predicted Reported | 205934 | 313504 | 508034 | 737161 | 1115710 |
|  |  | Predicted Total | 2813508 | 4382880 | 7219336 | 10557714 | 16066113 |
|  |  | Observed | 180298 | 275640 | 431719 | 584754 | 808306 |
|  |  | Underreporting Factor | 13.66 | 13.98 | 14.21 | 14.32 | 14.40 |
|  | Deaths | Predicted Reported | 7622 | 11837 | 19547 | 28840 | 44741 |
|  |  | Predicted Total | 32919 | 51921 | 86709 | 128523 | 199695 |
|  |  | Observed | 8053 | 10928 | 15316 | 19749 | 24903 |
|  |  | Underreporting Factor | 4.32 | 4.39 | 4.44 | 4.46 | 4.46 |

## 5 Simulations

Since the underlying truth is unknown in an actual study and the number of true latent infections are unobservable, we study the effect of selection bias and misclassification on the estimation of *R*_0_ and the predicted case counts via simulation studies where we know the true values. Each simulation is repeated 1000 times and average values/curves are reported.

### 5.1 Effect of Misclassification

We quantify the effect of incorporating false negative tests in our model by characterizing the differences in estimated *R*_0_ and predicted counts in various compartments (observed and unobserved) when we consider misclassification and when we do not. We first simulate counts from a model where the tests have false negatives and then estimate the quantities of interest using three models - one which considers the correct *f*, another which considers an incorrect *f* and the last one which considers *f* =0 or no false negatives. For this simulation we do not consider selection bias. We assume that all individuals are equally likely to be tested.

#### Generation Model

We generate the data using our 2 parameter Multinomial model with f = 0.3. The other parameters are fixed as in the data analysis for India: *N* = 1.341 billion, 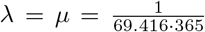, *α*_*p*_ = 0.5, *α*_*u*_ = 0.5, *β*_1_ = 0.6, *β*_2_ = 0.7, *δ*_1_ = 0.3, *δ*_2_ = 0.7, D_e_ = 5.2, D_R_ = 14, 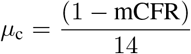 where mCFR = 0.054. We generate the data for a period of 101 days. The entire period was divided into five time periods which are days 1 − 10, 11 − 31, 32 − 50, 51 − 64, 65 − 101.

The values of *β* across the five periods are set at 0.8, 0.65, 0.4, 0.3, 0.3 and the corresponding values of r are set at 0.1, 0.2, 0.15, 0.15, 0.2. The chosen true values closely mimic the estimates for India from 15^th^ March to 23^rd^ June and the periods mimic the lockdowns in India.

#### Estimation Model

We choose the 2-parameter Multinomial model for estimation. We fit the model using the same parameters as in the model used for generating the data except *β*, r and f. We consider f = 0, 0.15 and 0.3 respectively for prediction in the 3 scenarios. The values of *β* and r are then estimated in each of the 3 scenarios for the 5 time periods. The entire process is repeated 1000 times. **Results:** <u>Estimation of</u> *<u>R</u>*<u>_0_</u>: The values of *R*_0_ for the five periods used to generate the data were 3.99, 3.65, 2.12, 1.59 and 1.69. The mean of predicted values of *R*_0_ for the model with *f* = 0 across the 1000 iterations were 3.64, 3.51, 1.97, 1.48 and 1.65 for the 5 periods while those for model with *f* = 0.15 were 3.52, 3.64, 2.01, 1.51 and 1.69 and for model with *f* = 0.3 were 3.83, 3.73, 2.04, 1.53 and 1.71. All the models have very low MRPE in predicting R_0_. The mean MRPE across 1000 simulations were 0.004, 0.004 and 0.001 for models with *f* = 0, 0.15 and 0.3 respectively. We will now look at predicted case count.

##### Prediction accuracy for total active case-counts

Figure 8 shows the variation of predicted values of total active cases (reported + false negatives+untested) across different models with varying rates of false negatives in a random instance of the simulation. We can observe that the model with *f* = 0.3 performs best in predicting the total active cases followed by the model with *f* = 0.15. As expected, the model which does not consider false negatives, i.e. with *f* =0 performs worst. When we calculate the mean of MRPE of the predicted number of reported active cases relative to that of the simulated true data across 1000 simulations, we note that the mean MRPE for the model with *f* = 0.3 is lowest with mean MRPE = 0.012 while the mean MRPE for the model with *f* =0 is highest with mean MRPE = 0.13. The mean MRPE for model with *f* = 0.15 lies in the middle with mean MRPE = 0.068. Thus we notice that there is a large gain in prediction accuracy if one incorporates the false negative rate of the test and it matches the true rate. We also note that the MRPE for reported active cases are considerably less. All the 3 models have a mean MRPE of 0.01 for reported active cases. This shows that different levels of misclassification only influences the estimates of total active cases substantially but not that of reported active cases. We see a similar trend in other counts as well. For cumulative cases, we have mean MRPE of 0.13, 0.066 and 0.012 for models with *f* = 0, 0.15 and 0.3 respectively. For total recoveries, we have mean MRPE of 0.12, 0.065 and 0.013 for models with *f* = 0, 0.15 and 0.3 respectively. Finally, for total deaths, we have MRPE of 0.06, 0.01 and 0.02 for models with *f* = 0, 0.15 and 0.3 respectively.

**Figure 8:**
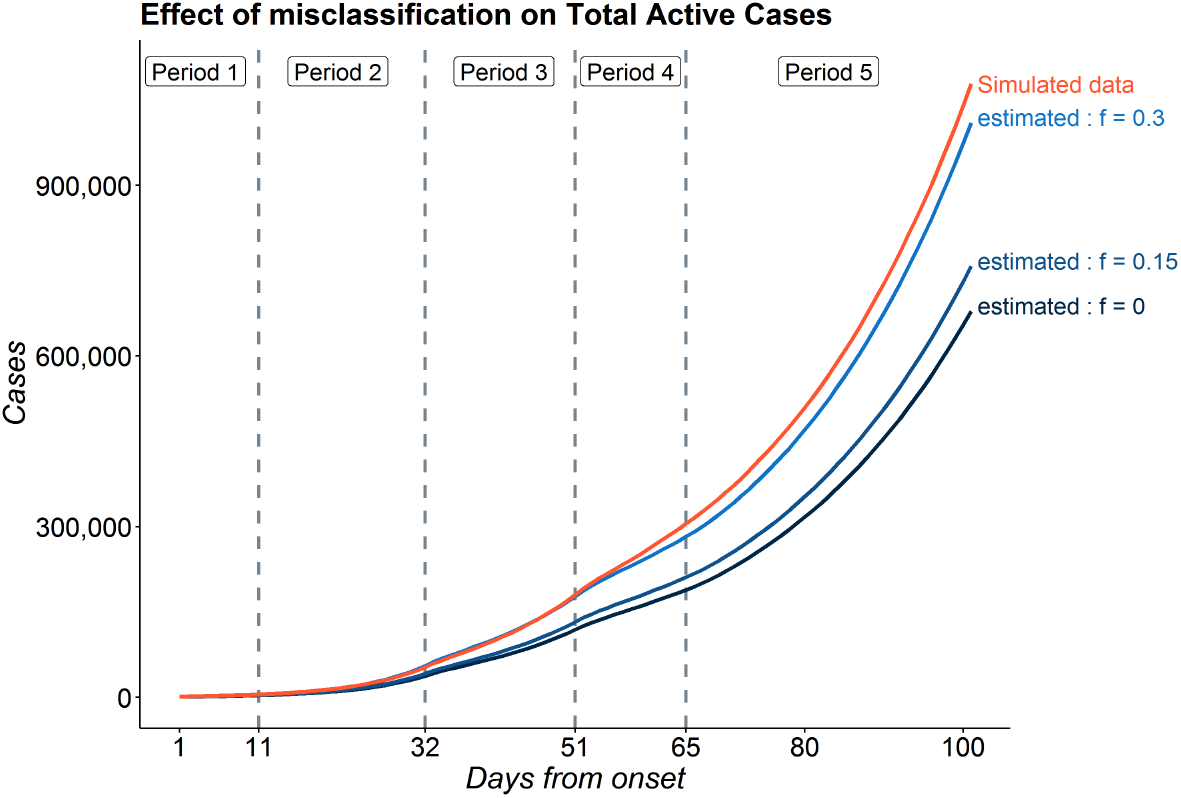
Variation of predictions with different rates of misclassification

### 5.2 Effect of Selection

In Section 3.3(Extension 3), we propose an extended model incorporating symptom-dependent testing. To study the effect of ignoring this testing mechanism, we generate data following this and estimate model parameters using a model that incorrectly ignores selection/testing(that is our misclassification model)

#### Generation Model

We generate data using Extension 3 model with most of the parameters same as the above previous simulation except : *α*_*u*_ = 0.7, 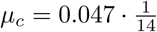, D_e_ = 5.2, D_r_ = 14/0.953. For Selection Model we have some extra parameters. They are set as *p*_0_ = (10^−6^, 10^−5^, 1 10^−6^ 10^−5^) and *p*_1_ = (0.02, 0.18, 0.8). As before, the data are generated for a period of 101 days with 5 periods 1 − 10, 11 − 31, 32 − 50, 51 − 64 and 65 − 101. The *β* values for the 5 periods were (0.6, 0.4, 0.3, 0.25, 0.2).

#### Estimation Model

Predictions are based on the Multinomial-2-parameter misclassification model, where the probability of being tested is assumed to be independent of symptoms with f = 0.3 (the simulation truth). Ignoring the misclassification will lead to even larger biases but we chose to decouple the effect of the two.

#### Results

##### Estimation of *R*_0_

The true values of *R*_0_ for the 5 periods used to generate the data were 2.22, 2.51, 1.89, 0.52 and 1.29. In presence of selection, we find that the estimated values of *R*_0_ differ substantially from the actual values. The means of estimated values across all the 1000 simulations for the 5 periods were 0.24, 2.40, 2.92, 2.56 and 2.10. Hence, we observe that the estimated value of *R*_0_ for the first period was much smaller compared to the actual value while the rest of the estimates were much better. The 2.5% quantiles of the estimates of *R*_0_ for the 5 periods across 1000 simulations were 0.02, 1.57, 2.64, 2.37 and 2.06 while the 97% quantiles were 0.78, 3.08, 3.18, 2.80 and 2.20.

##### Prediction accuracy of active case-counts

Figure 9 shows the predictions of Total and Reported Active cases in a random instance among the 1000 simulations. The blue band indicates the 95% CI of estimated counts in that particular simulation. The figure indicates that under selective testing, the counts predicted by the model may be very different than the true simulated data. The model incorporating misclassification and ignoring selection failed to capture the overall trend in the simulated data for the active cases (reported+unreported). This is more evident from the value of MRPE of the total active cases which has a mean of 0.56 with 0.32 and 1.54 as the 2.5% and 97.5% quantile respectively. While, for reported active cases, however, the misclassification model obtained fairly accurate predictions and successfully captured the trend in the data. The mean MRPE for the reported active cases is much lower than that of the total active cases. The mean MRPE for the reported active cases came out to be 0.085 with 0.044 and 0.15 as the 2.5% and 97.5% quantile respectively. Thus the effect of selection is more pronounced on the prediction of asymptomatic cases and what fraction of those cases get tested.

**Figure 9:**
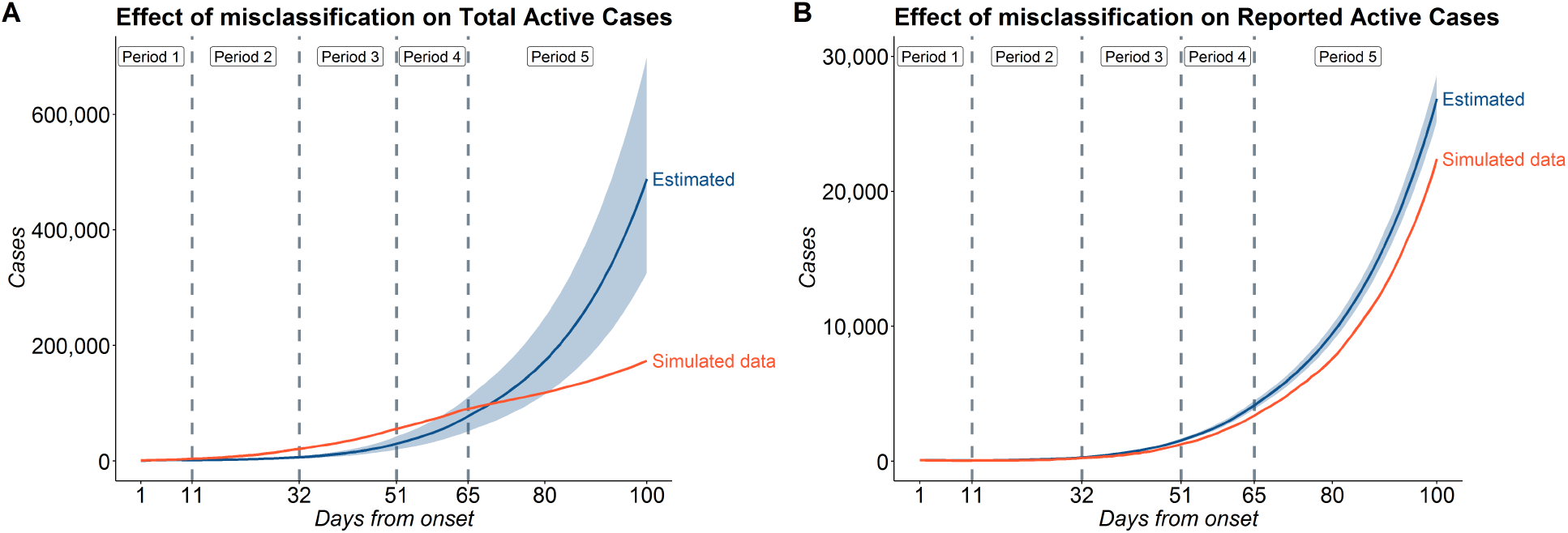
Effect of selection on (A) Total and (B) Reported Active Cases

This simulation demonstrates that selection bias has substantial impact on our estimates. Ignoring selection could lead to erroneous inference, particularly for predicting total number of true cases. Though we articulated this problem, we did not quite posit a solution in this paper as any solution will require accurate specification of the selection probabilities, which may not be known in practice or hard to estimate. The advantage of having a conceptual framework is that you can fix the selection probabilities and characterize this bias.

### 5.3 Effect of number of tests

In this section, we study the effect of increasing testing on the course of the pandemic. We expect that with increasing number of tests we have a better chance of identifying the infectious people, which might result in a faster end to the pandemic. To test this hypothesis, we use our Selection Model(Extension 3) to explore the population infection rates as a function of the number of available tests. **Generation Model:** We use the same Selection model for generating these data as in the previous section. To generate the data, we use five different scenarios where values of all the parameters except the number of tests is the same as in the previous simulation. The data are generated for a period of 1000 days with 5 periods 1 − 10, 11 − 31, 32 − 50, 51 − 64 and 65 − 1200. The values of *β* for the 5 periods were (0.6, 0.4, 0.3, 0.2, 0.05) for all the models. The only difference between the models is the number of tests. For the first scenario, we generated the number of tests such that the number of test increases exponentially from 20,000 on the first day to 1 million on the 1200^th^ day. We used the following equation for generating such a sequence :

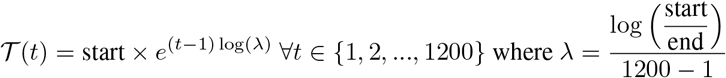

Let us denote the sequence of no of tests generated by 𝒯. Now, for the 5 scenarios, ated data using number of tests equal to 1, 2, 3, 4 and 5 times 𝒯 respectively. We repeat the process 1000 times and take analyze the mean predictions of total active cases.

#### Results

Figure 10 shows the mean number of total active cases across 1000 simulations as a function of time since pandemic onset. With a higher number of tests, the pandemic ends faster. We observe that the number of days before the first time the number of total active cases comes below the 1 million mark (after attaining the peak) for the models with number of tests = 3*𝒯* and 4*𝒯* is 1.14 and 1.06 times that of model with with number of tests = 5*𝒯*. For the models with number of tests =*𝒯* and 2*𝒯*, we note that cases do not come below the 1 million mark withing the 1200 day period.

**Figure 10:**
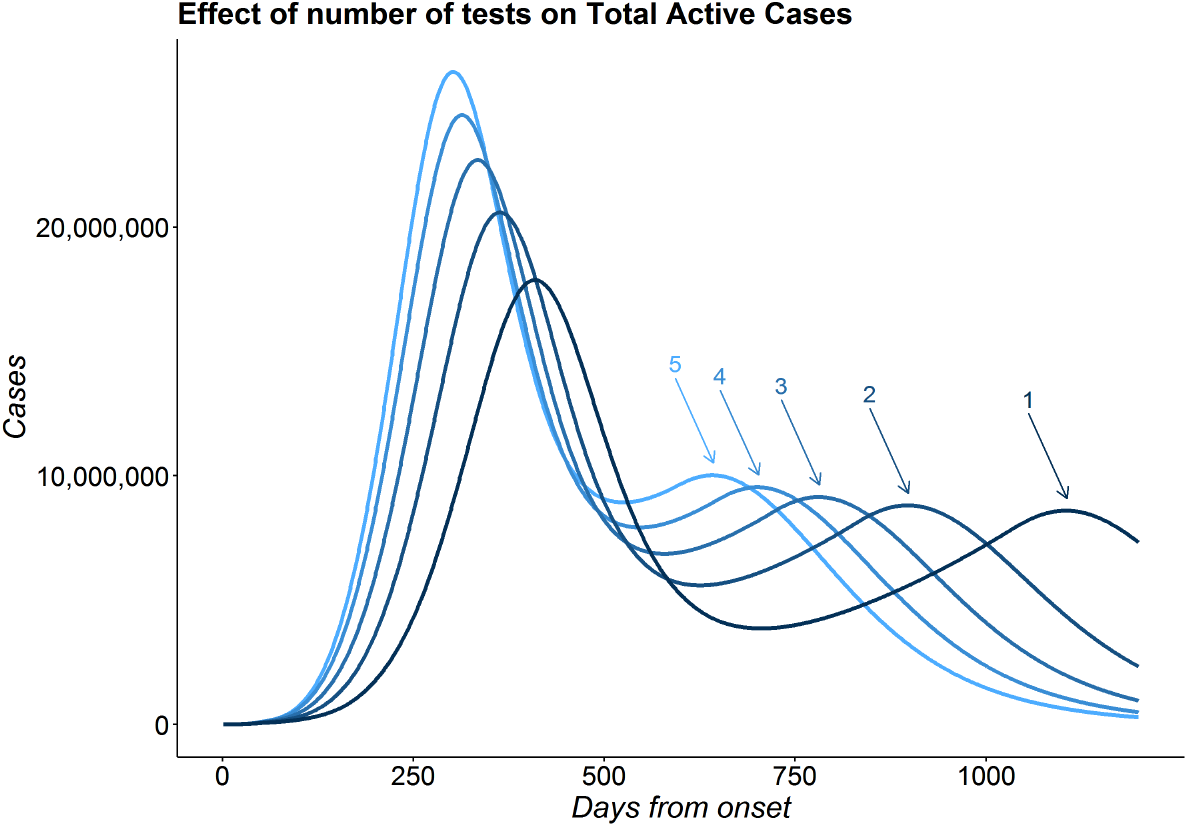
Effect of test : Plot showing the number of total active cases over time for different number of tests. The numbers above the arrows indicate the multiplication factor of number of tests

We also note that all the predictions in the 5 scenarios predict the existence of a 2^nd^ peak. This is a proof of concept illustration with our formulation. We further observe that with higher number of tests, the gap between the first and second peak becomes smaller. Here, for the models with number of tests equal to 2, 3, 4 and 5 times 𝒯, we have the gap between the 2 peaks as 0.77, 0.64, 0.56 and 0.49 times that of model with number of tests equal to 𝒯. We should note that while the actual values presented in this simulation do not bear any resemblance with those of any country or state, the relative orders and findings convey important insights regarding effect of the number of tests on the number of infections.

## 6 Sensitivity Analysis

We now refer back to our simulation studies in Section 5.1. Since we fixed many parameters in the SEIR-fansy model and only estimate *β* and r, we consider an exhaustive framework of sensitivity analysis focusing on certain key inputs listed below.

1. **E**_**0**_ : We have tried 4 different values of *E*_0_ and check how the estimates of *R*_0_ and Reported Active cases vary across different values of *E*_0_. We have taken *E*_0_ = 1, 2, 3 and 4 times (*U*_0_ + *P*_0_ + *F*_0_) in the 4 models for sensitivity analysis.
2. *α*_**U**_ : The value of *α*_*U*_ had been taken as 0.5 in the main analysis. We also assumed *α*_*P*_ = 0.5. So, we try 4 different values of *α*_*U*_ here which are *α*_*U*_ = 0.3, 0.5, 0.7 and 1.
3. **D**_**e**_ : We have taken D_*e*_ = 5.2 days which is same as the incubation period. Here we have considered 3 values of D_*e*_ for sensitivity analysis. They are D_*e*_ = 6.4, 5.2 and 4.1.
4. **k** : For Multinomial Symptoms model, one important parameters is *k* which is the ratio of probability of a mildly In our main analysis, we assumed *k* = 4. Here we have tried 4 different values of *k* : *k* = 3, 4, 5 and 6 and look at the different estimates.

We examine the estimates of the basic reproduction number for the different periods as well as the total active number of cases for the different periods. The basic findings from the sensitivity analyses are summarized as follows:

- As expected, the predictions for the Reported Active cases (P) remains same.
- The estimates for *R*_0_ mainly differs in the first (and slightly in the second period) but are almost same for the later stages of the pandemic in the different models.
- For the untested cases, in some of the cases there are substantial deviations from the generated cohort for a particular parameter. The total number of active cases, which includes both the unreported and the reported cases, varies substantially with different parameter values. This shows that estimation of unreported cases is quite sensitive with respect to different choices for the parameter values. In particular, we see the highest variation with different values of *E*_0_ and the least variation for *D*_*e*_.

For detailed discussion on the sensitivity analysis, please refer to the Section 12 of the Supplementary Methods.

## 7 Conclusion

In this paper we have considered a mathematical framework for incorporating selection bias and misclassification in a traditional SEIR model and illustrated the methods with reported COVID case, recovery and death counts in India from April 1-August 31. The study has several limitations.

- Like all compartmental models, our model also assumes a structure for the dynamics of disease transmission. When the model assumptions do not hold, resulting estimates may be inaccurate.
- Unlike other compartmental models, we do not assume homogeneous mixing of the population. We assume different rates of infection from individuals in *U, P* and *F*. However, we assume that the probability of contracting the virus is the same for all individuals in a particular compartment. This does not reflect the realistic scenario where contact happens within individualized local networks. When this assumption is not tenable, it is better to apply the model to smaller sub-populations and add them up to obtain prediction for the larger population. Alternatively, one could decompose each compartment into sub-compartments based on age-sex-job specific contact network and impose a hierarchical structure.
- Our model also assumes that the values of *β* and *r* remain constant over chunks of time and that other parameters remain constant through the entire course of the pandemic. However, such an assumption might not be true in real life. The value of these parameters vary over time and change almost everyday. Nevertheless, when public health measures remain invariant and under the absence of any disruptive event like a large superspreader event, the variation in values in minimal and hence, we can assume them to be constant.
- In this paper, we have mainly focused on the accuracy of predicted reported cases with less attention given to prediction of reported recoveries and deaths. This is mainly because of the model structure and the dichotomy between what parameters are estimated and what parameters are assumed to be known. In the Multinomial-2-parameter model, we estimated the parameters *β* and *r* that are directly related to the active number of cases. If we are precise in terms of predicting this count, our predictions of all other compartment counts improve given that our assumptions regarding the other fixed parameters are compatible with the data. We have chosen reasonable values of the parameters related to Recoveries and Deaths. The success of estimating the counts in *R* and *D* relies largely on our assumptions and our estimation of active case counts. There is an extensive body of work starting from the death data [9] that adopts the reverse strategy, and future comparison of our method with this genre of work is warranted.
- For predicting future cases, we assume values of *β* and *r* remain the same as in the last period. This might not be true with changes in human behavior and public health measures to control the virus. One possible solution is to replace *β* by *β π*(t) as done by Ray et al. [17]. Here, *π*(t) is a time varying intervention modifier which takes values in [0, 1] for stricter lockdown relative to the last period in the training data. Similarly, it takes values greater than 1 for less stringent lockdown relative to the last period or when lockdown is lifted.
- Our treatment of the selection component is largely incomplete as it relies on knowing/estimating a selection model for testing. While in some regional levels this may be well-characterized, it will be hard to know this at scale for an entire country.

In spite of all these limitations, our SEIR-fansy method has some major strengths.

- Our model accounts for misclassification in the form of false negatives. We have shown that misclassification has a substantial effect on the number of total cases (reported + unreported). Kucirka et al. [14] show that the sensitivity of RT-PCR tests for COVID-19 can be as low as 0.7, which translates to a 30% false negative rate (FNR). We show that accounting for 30% FNR can result in the estimates of total active cases to be about 50% higher than estimates under perfect testing.
- Our model also considers a compartment for Untested individuals, which is very important when modelling COVID-19. Recent serological surveys in Delhi [19, 20, 3] have shown that only 2.4% of the cases are being reported (Underreporting factor of about 42). In our model *U* (Untested) and *F* (False Negatives) together represent the number of unreported active cases while *RU* and *DU* represent the number of unreported recovered and deaths respectively.
- We have derived a neat expression of the *R*_0_ under complex design and data quality issues, which provides intuition regarding the influence of these processes on inference from a compartmental model.
- Unlike the conventional SEIR model, we use Bayesian modelling, which enables us to quantify uncertainties in model parameters. We have presented 95% credible intervals of model parameters as well as average posterior compartmental counts.
- Our selection framework allows us to fix the selection probabilities and study their effect on the estimates and quantify the resultant bias. This can serve as an excellent sensitivity analysis tool.
- We have provided open-source code for implementing our method.

The paper has both theoretical and practical significance. The SEIR-fansy method proposes a comprehensive framework to conceptualize incorporation of false negatives and selective testing in a SEIR model, which has broader methodological significance for modeling any infectious disease transmission where testing/diagnostic strategies are imperfect. We have fit our model on India and two of its states, Delhi and Maharashtra. We estimate that the **underreporting factors** for cumulative cases in India, Delhi, and Maharashtra are approximately **21, 54**, and **14** respectively, while that for cumulative deaths are **6, 12**, and **4** respectively as of 1^st^ September. The implications of these results are both positive and negative. While a large number of unreported cases indicates failure in detecting a large number of covert infections, it also implies a considerably lower infection fatality rate (IFR) (which is defined by total no of deaths divided by the total number of cases). The estimates of **IFR** for India, Delhi and Maharashtra as of 1^st^ September are **0**.**88%, 0**.**91%** and **1**.**24%** respectively. The WHO [28] reports the value of IFR for COVID-19 to be between **0**.**5** to **1%**. The case-fatality rate (CFR) for India, Delhi, and Maharashtra are 1.74%, 2.46%, and 3.08% respectively. Thus the estimates of IFR are considerably lower than CFR which show that the virus is less lethal than it initially appeared.

Apart from these results in the context of India, we have also conducted an extensive simulation study to characterize the effects of misclassification, selection and number of tests on case counts and model parameters. We observe that misclassification alone does not have a substantial effect on estimates of *R*_0_ but has a strong effect on projected counts. Selection bias, on the other hand, has a strong effect on estimates of *R*_0_ and case counts. However, the effect of selection on estimates of *R*_0_ is not as pronounced as on case counts. We also perform a simulation study to understand the effect of number of tests on pandemic duration. We observe that with higher number of tests, the pandemic is declining at a faster rate, which shows the importance of increasing number of tests. Finally, we have performed elaborate sensitivity analyses for different parameters in our models, indicating that the estimates of *R*_0_ are relatively robust with respect to different choices of parameters but that estimates of the number of cases are sensitive to these choices. In particular, the estimated counts for the latent/unobserved compartments heavily rely on these assumptions.

## Data Availability

Code for R package available: https://github.com/umich-biostatistics/SEIRfansy

## 8 Acknowledgement

This research is supported by the Michigan Institute of Data Science (MIDAS), Precision Health Initiative and Rogel Scholar Fund at the University of Michigan. The research of BM are supported by NSF DMS 1712933 and CA 046592.

## Supplementary Materials

### 1 Differential Equations for Non-Instantaneous Testing

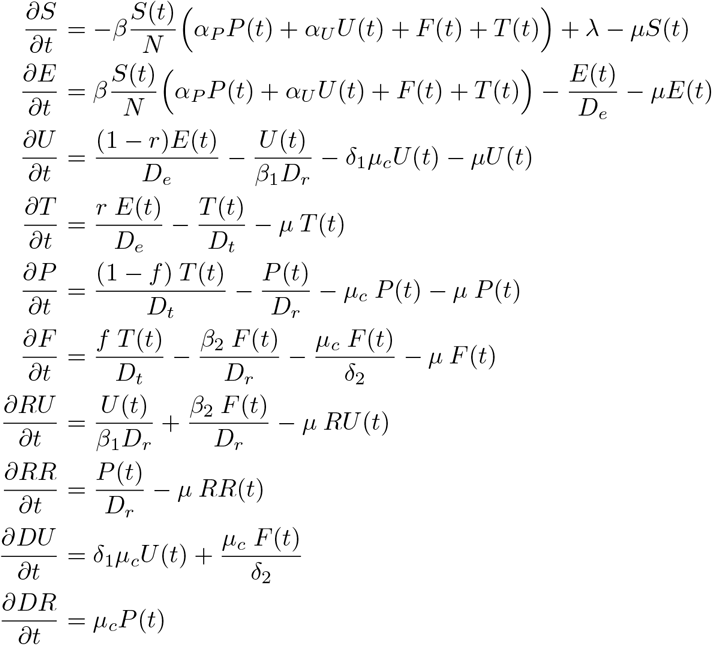

### 2 Calculation of *R*_0_ for Misclassification Model

We calculate the basic reproduction number *R*_0_ using the **The Next Generation Matrix Method** as described by van den Driessche [3]. Suppose the whole population is divided into n compartments in which there are *m* < *n* infected compartments. Let *x*_*i*_,*i* = 1, 2,.. , *m* be the number of infected individuals in the *i*^th^ infected compartment at time *t*. Now, the epidemic model is:

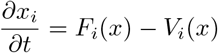

Here, 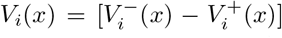, where 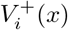 represents the rate of transfer of individuals into compartment *i* from all other components containing individuals infected with the disease (here *E, U, P* and *F*) and where 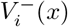 represents the rate of transfer of individuals out of compartment *i*. Here, *F*_*i*_(*x*) represents the rate of appearance of new infections in compartment *i*. Let *x*_0_ denote the disease free equilibrium. Now *ℱ* and *𝒱* are *m*×*m* matrices such that :

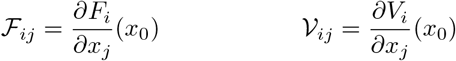

Now, *ℱ 𝒱*^−1^ is called the **Next Generation Matrix**. The basic reproduction number *R*_0_ is calculated by the spectral radius or the largest eigenvalue of *ℱ𝒱*^−1^. For our case,

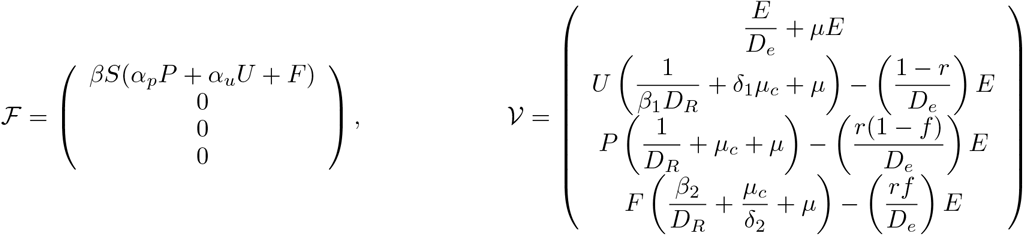

Now, we calculate the jacobian of *ℱ* and *𝒱* at the Disease Free Equilibrium (DFE).

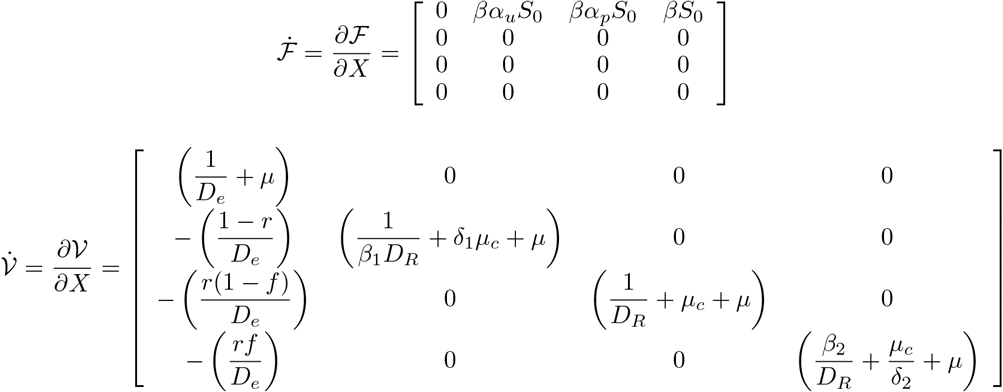

Now, we need to find the inverse of 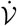. Since it is a lower triangular matrix, it is easy to find the inverse.

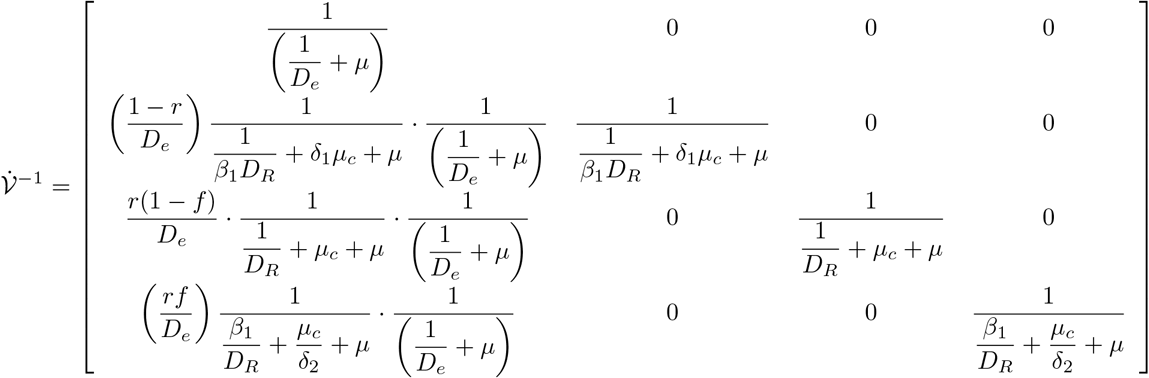

Now, we multiply 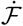 and 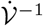. The spectral radius of 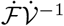 gives the basic reproduction number. Note that the matrix 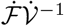 has only one non-zero row, which is the first one. All other rows of 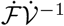 are 0. Hence, the spectral radius is given by 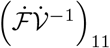 (i.e., the (1, 1)^*th*^ element of 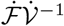). So,

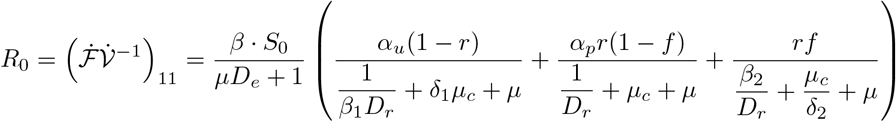

### 3 Motivation Behind the 3-parameter Multinomial Model

We can observe from figure (S1) that mCFR varies widely across countries and also across time. Now note that while countries like Belgium, USA, Italy, Spain have very high mCFR, India and Russia have comparatively much lower mCFR. Also, we observe that initially most countries experience high mCFR and it gradually settles to a comparatively lower value in most countries as the case counts and recoveries rise. This supports modeling mCFR as a time-varying quantity.

**Figure S1:**
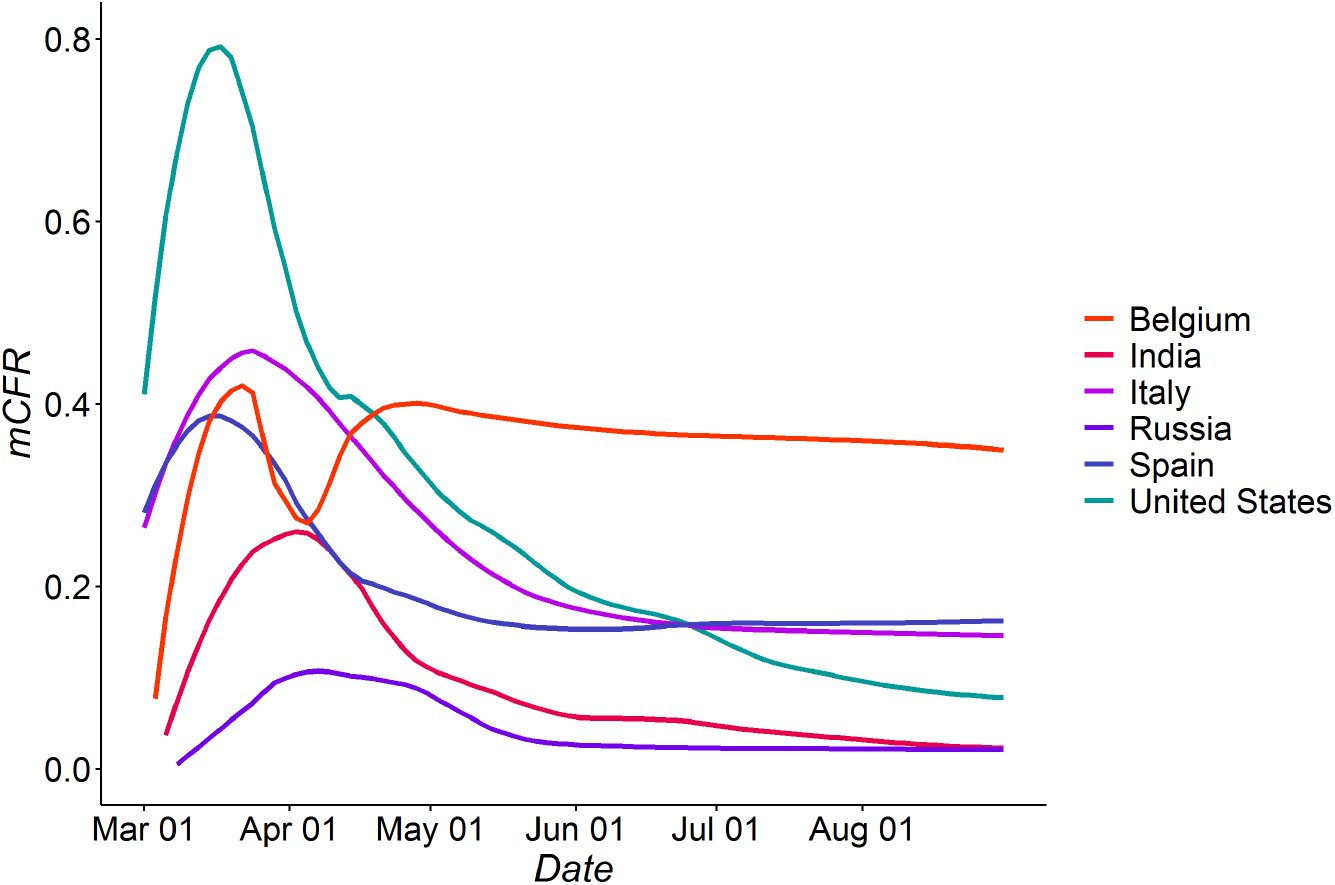
Variation of mCFR with time

### 4 Transmission Dynamics Diagram for Symptoms Model

**Figure S2:**
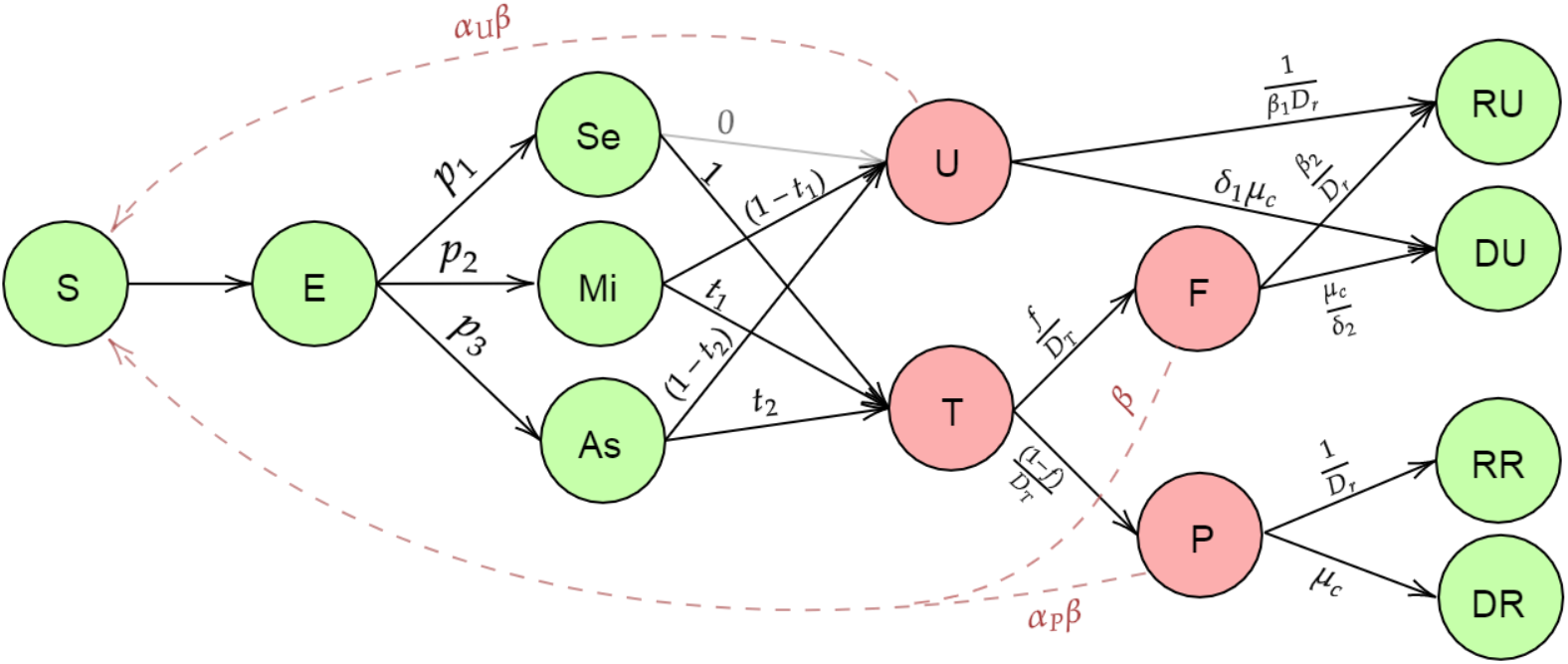
Misclassification Model with Symptoms

The only difference in this model from the Multinomial 2-parameter model is that, from the Exposed Node, an infected person can enter into one of three possible nodes: Severe Symptomatic Infectious(Se), Mild Symptomatic Infectious(Mi) and Asymptomatic Infectious(As) with probabilities *p*_1_, *p*_2_, and *p*_3_ respectively. Now, we assume people with severe symptoms (people in *Se*) are tested with probability 1. While the *Mi* people and the *As* people are tested with probabilities *t*_1_ and *t*_2_.

### 5 Misclassification model - complete distributional assumptions

In the main paper, we have given the distribution of observed nodes given the other nodes and parameters. Here, we describe the distribution of the latent nodes also. After getting the estimates of the parameters using MCMC, we want to obtain model-based forecasts. In order to predict the future counts, we use the following multinomial random sampling strategy:

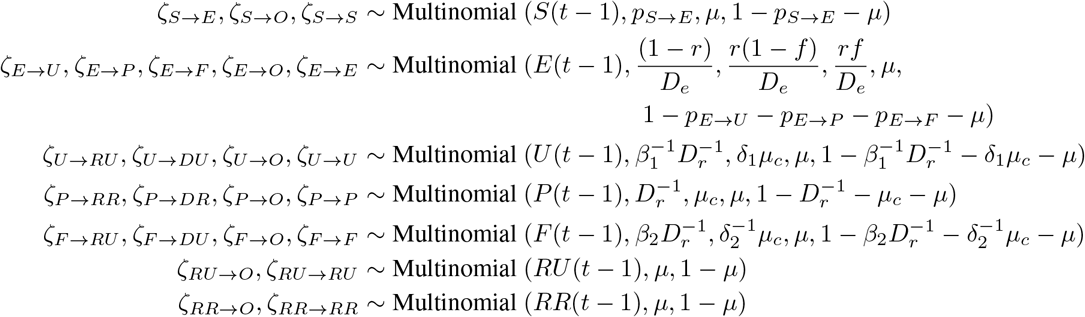

where *ζ*_*X*→*Y*_ denotes the number of individuals moving from compartment *X* to compartment *Y* at time *t. ζ*_*X*→0_ denotes the number of individuals in compartment *X* that die at time *t*. The counts in each compartment at time *t* are given by,

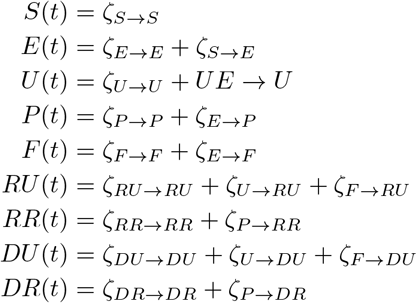

Given the parameters and the counts at time (*t* − 1), we obtain the predicted counts for time *t*. Using this approach, we obtain the posterior means of the future predicted counts at each of the 9 compartments using the MCMC estimated parameters. For the purpose of future prediction beyond the training period, we use the parameter estimates from the last time period.

### 6 Differential Equations for the Selection Model

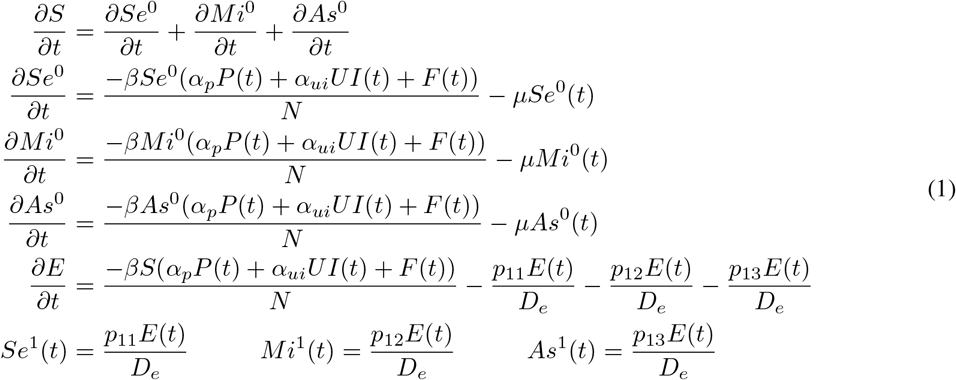

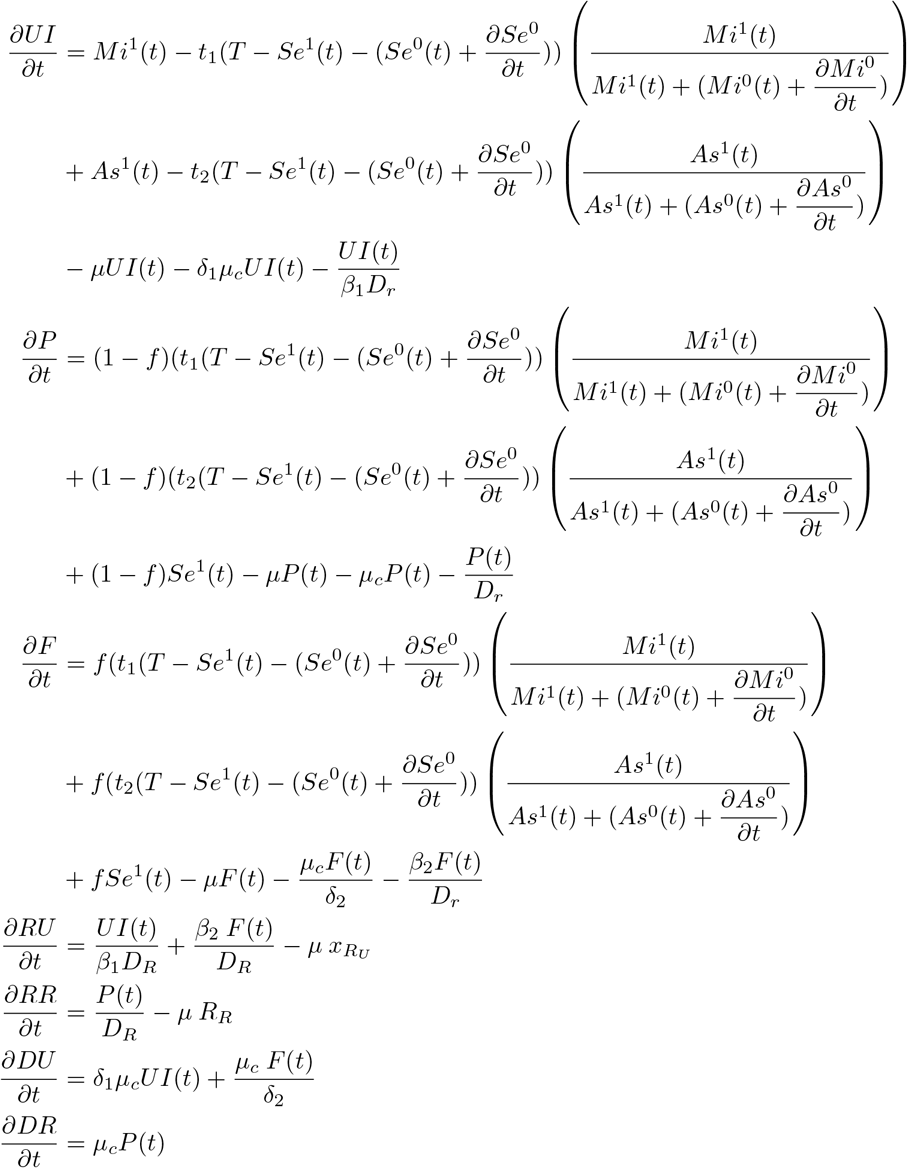

### 7 Selection Model : Complete Distributional Assumptions

To generate data using the test model, we perform the following steps.

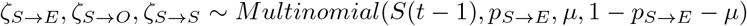

Now, we assume the probability of an individual being severely symptomatic, mildly symptomatic or asymptomatic given he/she is susceptible is given by the probability vector **p**_**0**_ = (*p*_01_, *p*_02_, *p*_03_). The probability for an infected individual is given by **p**_**1**_ = (*p*_11_, *p*_12_, *p*_13_). To obtain the number of individuals in the groups *Se*^0^,*Mi*^0^, *andAs*^0^, we assume that the outgoing individuals from the susceptible group follow the distribution given by **p**_**0**_.

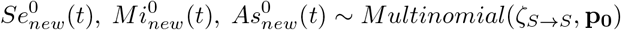

Now, from our assumption that the individuals in *E* follow the distribution given by **p**_**1**_, we can write,

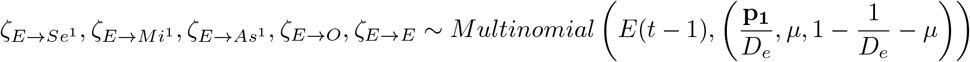

Recall, we assume that all individuals with severe symptoms are tested provided adequate tests are available. This implies

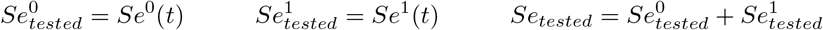

In the case when number of test *T* (*t*) is less than that of severe individuals, we assume that the number of tested *Se*^1^ and *Se*^0^ individuals is proportional to their respective counts.

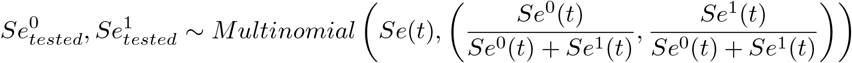

If the total number of remaining tests is greater than or equal to the number of mild and asymptomatic individuals, then all of them are tested i.e :

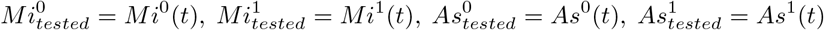

If number of tests are not adequate for all the mild symptomatic and asymptomatic people to be tested, then the remaining tests (after testing the severe symptomatic people) are distributed among the mildly symptomatic and asymptomatic individuals in the ratio *t*_1_ : *t*_2_.

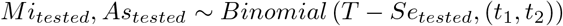

As we did in the case of severely symptomatic, we allocate the tests among infected and uninfected mildly symptomatic (and also asymptomatic) individuals randomly.

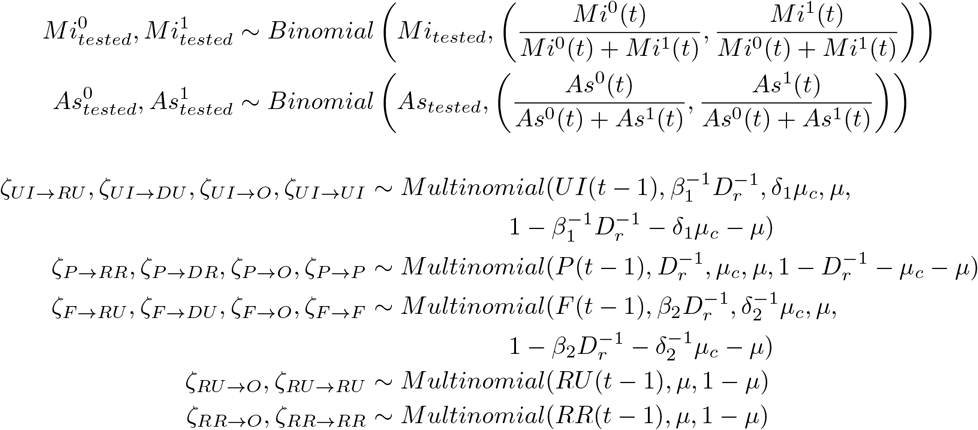

We also assume the false negative probability = *f*. The numbers of new individuals to P and F states are given by :

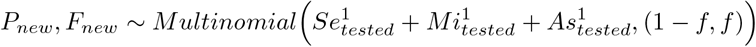

Finally we write the number of people in each state at time *t* as follows :

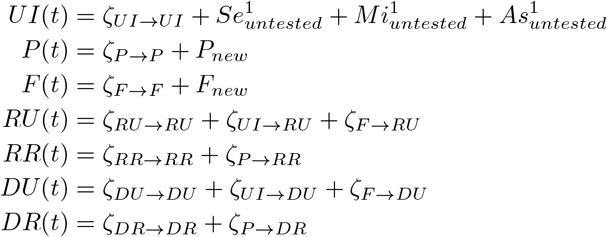

**Figure S3:**
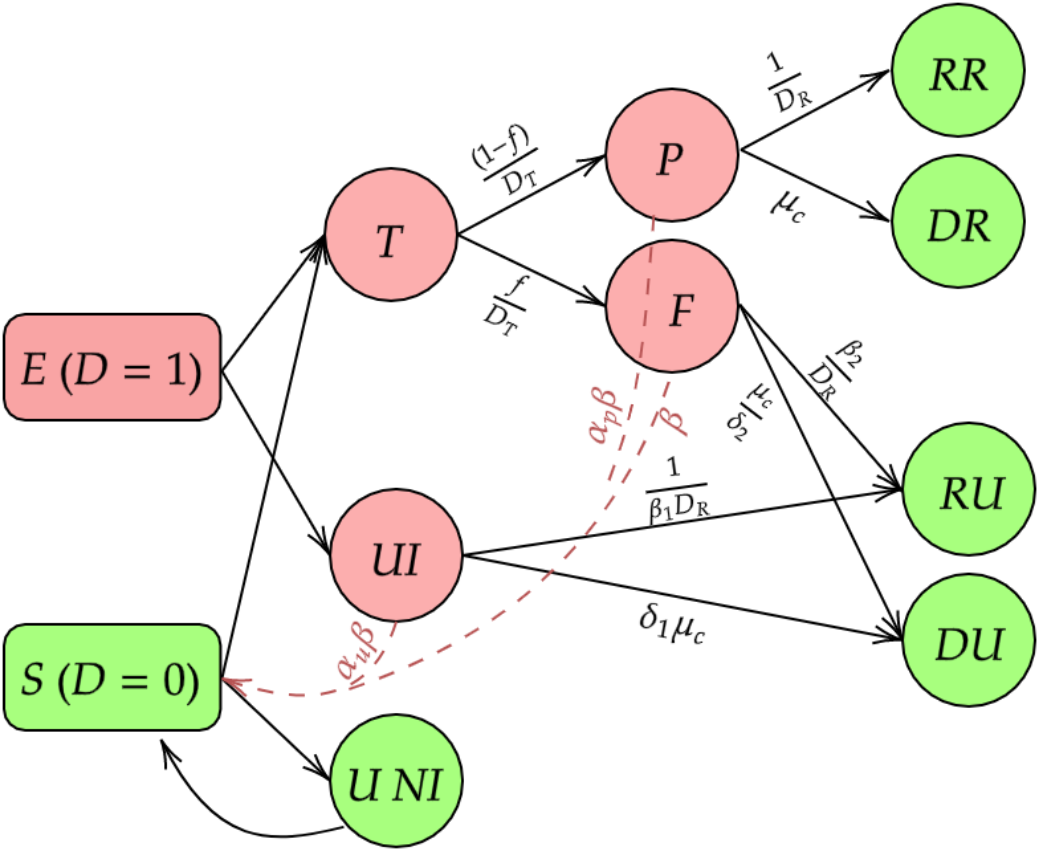
Special case of Selection Model : Uniform testing

### 8 Special case of Selection Model : Uniform testing

To understand the effect of selection bias on *R*_0_, we consider a special case of the Selection model where we assume uniform testing. Here, uniform testing means tests are offered independently of symptoms. The model is represented diagrammatically in Figure (S3).

The transmission dynamics of this model are very similar to the Selection model. We provide the differential equations describing the dynamics of the nodes *S, E, UI, P* and *F*. The rest of the nodes (*RU, RR, DU* and *DR*) have differential equations exactly same as in Selection Model.

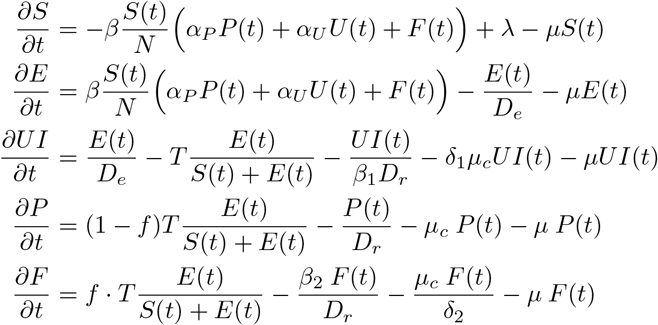

Now that we have all the differential equations governing the dynamics of this model, we calculate the basic reproduction number using Next Generation Matrix method [3]. Using calculations similar to what we did for the Misclassification model, we arrive at the following expression of *R*_0_ for Uniform testing model.

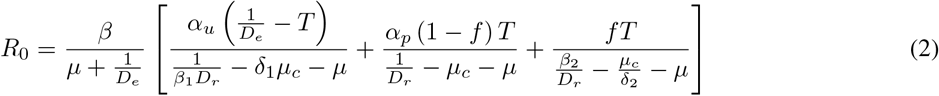

### 9 Real Data Analysis for India

#### 9.1 Table for Initial Values of the Different Compartments for India

**Table S1:** Initial Values of the Different Compartments

| Variable | Value | Justification |
| --- | --- | --- |
| S(0) | 1340933446 | $N - (E(0) + U(0) + P(0) + F(0) + RU(0) + RR(0) + DU(0) + DR(0))$ ($N = 1341$ million) |
| E(0) | 48780 | Thrice the number of current infected |
| U(0) | 13821 | $\frac{1-r}{r} (P(0) + F(0))$ |
| P(0) | 1829 | Reported current infected on 1<sup>st</sup> April |
| F(0) | 610 | $\frac{f}{1-f} P(0)$ |
| RU(0) | 958 | $\left(\frac{1-r}{r} + \frac{f}{1-f}\right) RR(0)$ |
| RR(0) | 169 | Reported recovered on 1<sup>st</sup> April |
| DU(0) | 329 | $\left(\frac{1-r}{r} + \frac{f}{1-f}\right) DR(0)$ |
| DR(0) | 58 | Reported deceased on 1<sup>st</sup> April |

Now for India, we have fitted data from 1^st^ April to 30^st^ June. So for our prediction, we need the counts of the different compartments on the initial date, that is on 1^st^ April. So the table S1 presents the counts of the compartments for India on 1^st^ April.

#### 9.2 Additional Plots for India

We have done our estimation using the MCMC Metropolis Method and predicted the counts for the different compartments by using the posterior means conditional on the estimated parameters. So the large number of iterations of MCMC provide a 95% credible interval for the parameters as well as for the predictions of the compartments. So the following figure shows the credible regions for the Reported Active, False negative active and Untested Active cases.

**Figure S4:**
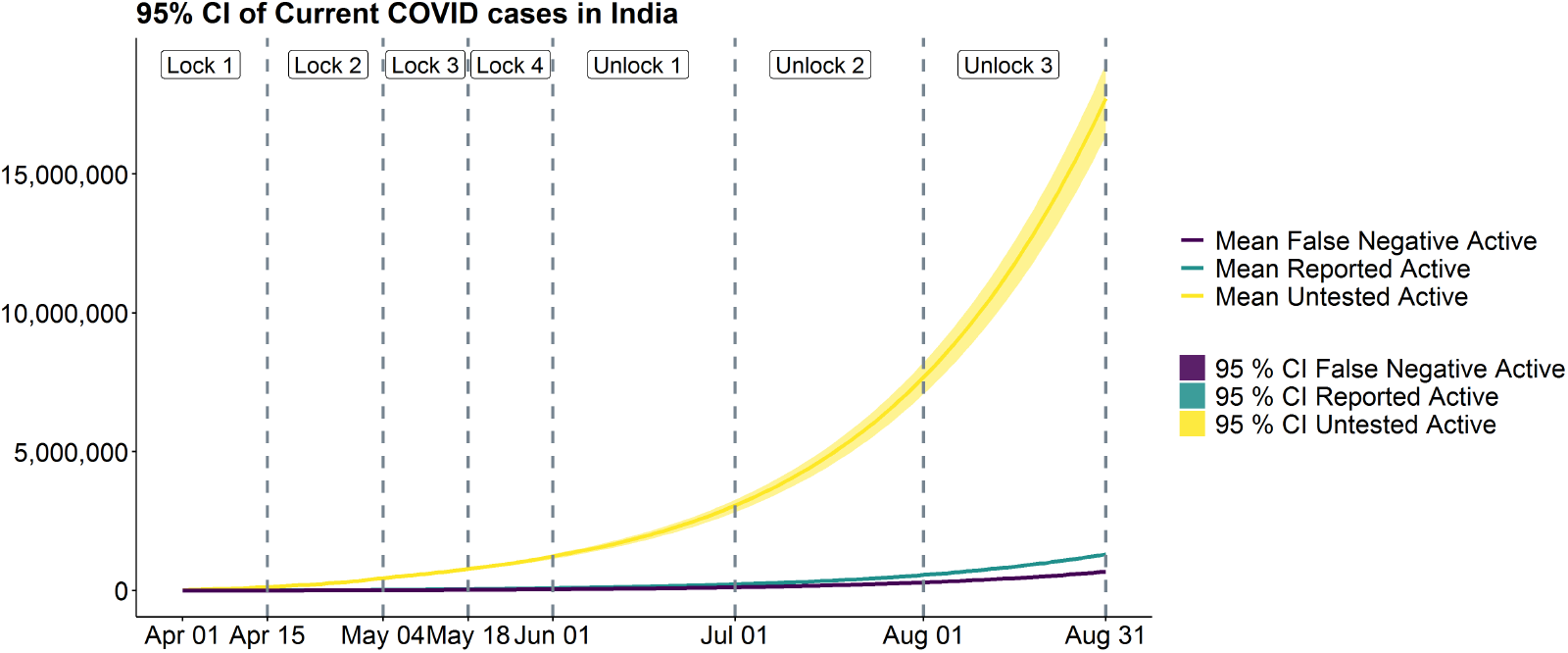
95% Credible Intervals of estimates of Current Active Cases in India

Figure (S4), shows the 95% CI’s of the estimates of Current Active cases in India from 1^st^ April to 31^st^ August. Here, we have fit the model using the data from 1^st^ April to 30^th^ June dividing the training period into 5 parts as described earlier. The estimates of *β* and *r* for the last period (1^st^ June to 30^th^ June) was used to predict the cases from 1^st^ July to 31^th^ August. As it is easily visible from Figure (S4), the CI’s for estimates of untested cases is the highest. This is expected due to the much higher estimated number of untested cases than tested positives or false negatives.

### 10 Additional Plots for Maharashtra and Delhi

**Figure S5:**
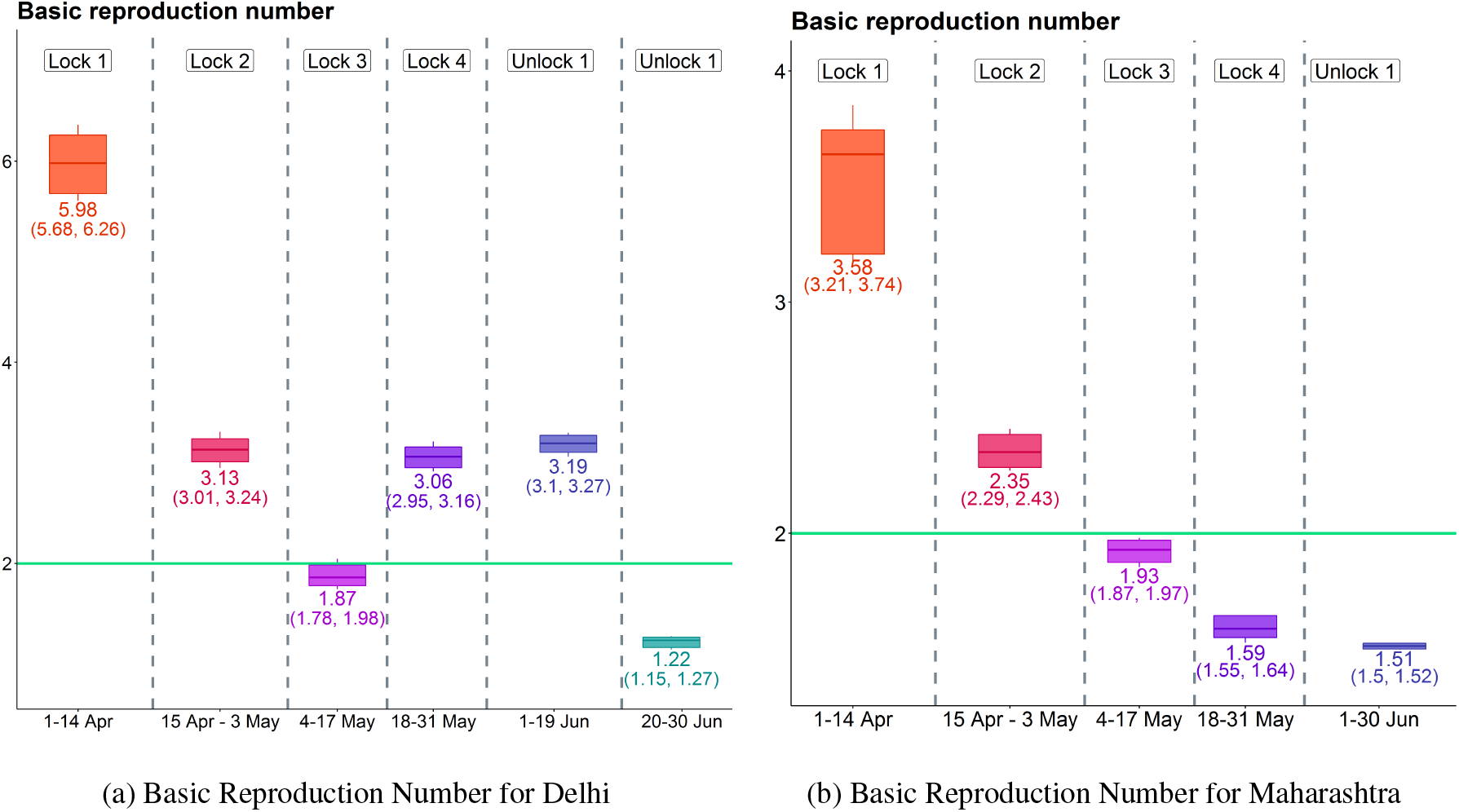
Basic reproduction number of Delhi and Maharashtra

From Figure (S5b), we observe that the estimates of basic reproduction number for Maharashtra have been strictly decreasing throughout the training period. The value of *R*_0_ started at 3.58 in the first 2 weeks of April and eventually dropped to 1.51 in June. The value of *R*_0_ dropped below 2 first time in lockdown 3 which was from 4^th^ to 17^th^ May. From figure (S5a), we note that the basic reproduction number was quite high (> 3) in 1^st^, 2^nd^, 4^th^ and 5^th^ periods and it decreased to 1.22 in the 6^th^ period.

### 11 Results for Simulations - Effect of Misclassification

In the main paper we have shown the effect of misclassification on number of total active cases. We concluded that the effect of misclassification on total active cases was substantial, but it was negligible on reported active cases. Here, we provide the mean estimates of *R*_0_ obtained by the 3 different models with 3 different false negative rates *f* = 0, 0.15 and 0.3.

**Table S2:** Effect on Basic Reproduction Number

|  | Basic Reproduction Number |  |  |  |  | MRE |  |  |
| --- | --- | --- | --- | --- | --- | --- | --- | --- |
| | $R_{01}$ | $R_{02}$ | $R_{03}$ | $R_{04}$ | $R_{05}$ | Lower C.I | Mean | Upper C.I |
| Actual | 3.99 | 3.65 | 2.12 | 1.59 | 1.69 | - | - | - |
| Predicted Using $f = 0$ | 3.64 | 3.51 | 1.97 | 1.48 | 1.65 | 0.0036 | 0.0041 | 0.0045 |
| Predicted Using $f = 0.15$ | 3.52 | 3.64 | 2.01 | 1.51 | 1.69 | 0.0035 | 0.004 | 0.0044 |
| Predicted Using $f = 0.3$ | 3.83 | 3.73 | 2.04 | 1.53 | 1.71 | 0.0009 | 0.0012 | 0.0015 |

It is quite evident from the table S2 that the *R*_0_ is quite robust with the change of the value of false negative rate (*f*). Under all the false negative rates, the estimation of *R*_0_ is quite accurate which is evident from the MRE provided in the table S2.

### 12 Sensitivity Analysis

Since we have not estimated the values of quite a few parameters, a sensitivity analysis is necessary. Now, as doing sensitivity analysis of all the parameters and initial values is impractical, we will do sensitivity analysis for the following parameters only.

1. **E**_**0**_ : The initial value of Exposed had been chosen as 3 times the sum of initial values of Untested, Confirmed and False Negative cases. Such a choice might seem arbitrary. Hence, we try 4 different values of *E*_0_ and check how the estimates of *R*_0_ and Current Active cases vary across different values of *E*_0_.
2. *α*_**U**_ : The value of *α*_*U*_ had been taken as 0.5 in the main analysis. We also assumed *α*_*P*_ = 0.5. So, we effectively assumed that the rate of transmission of disease by untested and tested positive individuals was same. Some things to consider when choosing the value of *α*_*U*_ and *α*_*P*_ were that individuals who were tested positive are quarantined and/or hospitalized reducing their rate of transmitting the disease. And untested cases are predominantly asymptomatic cases whose rate of spreading the virus is much less than symptomatic cases. So, we have *α*_*U*_ < 1 and *α*_*P*_ < 1. However, we do not know if *α*_*U*_ > *α*_*P*_ or *α*_*U*_ < *α*_*P*_. So we try 4 different values of *α*_*U*_ here which are *α*_*U*_ = 0.3, 0.5, 0.7 and 1.
3. **D**_**e**_ : We stated in the beginning of this paper that we have assumed the Incubation period equals the Latency period (= *D*_*e*_). We have taken *D*_*e*_ = 5.2 days following the results by Lauer et al. [2]. However research by other groups suggest different values of incubation period like 6.4 days by Becker et al. [1] etc. So we consider 3 values of *D*_*e*_ for sensitivity analysis. They are *D*_*e*_ = 6.4, 5.2 and 4.1 (lower limit of 95% CI of estimates of incubation period by Lauer et al. [2])
4. **k** : For Multinomial Symptoms model, one important parameters is *k* which is the ratio of probability of a mildly symptomatic person being tested to that of an asymptomatic person being tested. Since the probability of testing is higher for a mildly symptomatic person than an asymptomatic person, so *k* > 1. In our main analysis, we assumed *k* = 4. The choice of *k* was not supported by any data. So, we try 4 different values of *k* : *k* = 3, 4, 5 and 6 and look at the different estimates.

#### 12.1 Effect of initial value of Exposed

We start with the initial value of Exposed individuals. Throughout our analysis we have assumed that the number of exposed individuals on the starting day i.e. 1^*st*^ April was thrice the number of total expected infected up to that day. So we check how much our estimates vary if we vary the starting value of Exposed (*E*_0_). So, we use 4 starting values for *E*_0_:

1. *E*_0_ = *U*_0_ + *P*_0_ + *F*_0_
2. *E*_0_ = 2(*U*_0_ + *P*_0_ + *F*_0_)
3. *E*_0_ = 3(*U*_0_ + *P*_0_ + *F*_0_)
4. *E*_0_ = 4(*U*_0_ + *P*_0_ + *F*_0_)

**Figure S6:**
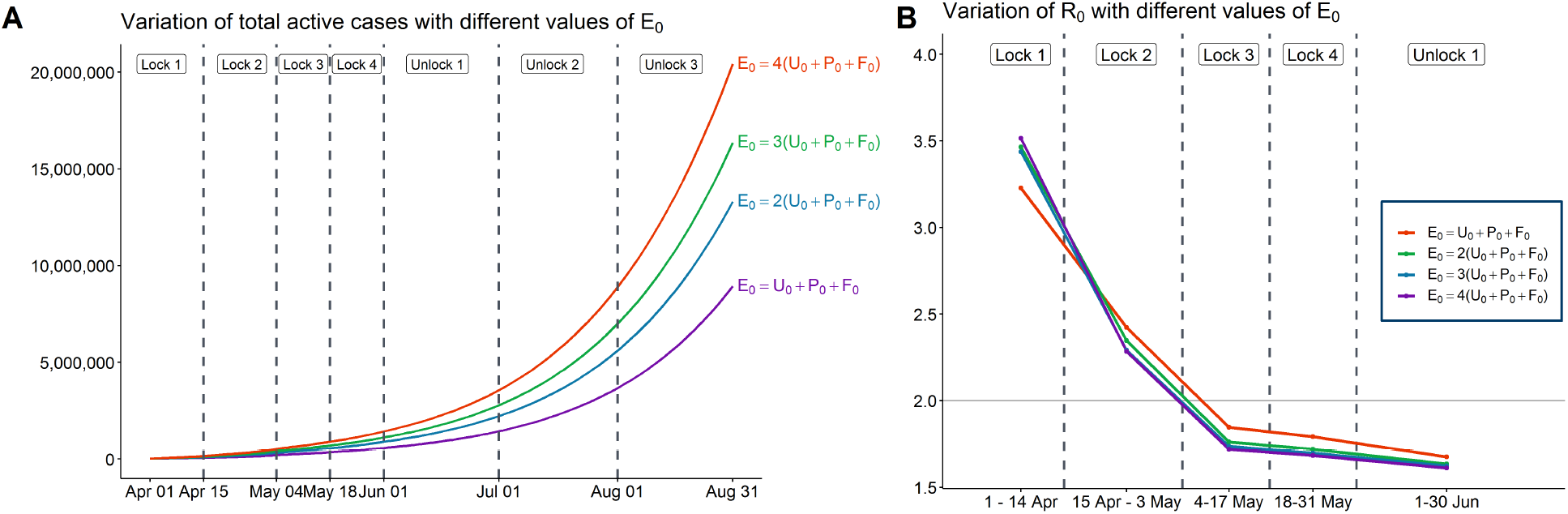
Variation of estimates of *R*_0_ with different values of *E*_0_

We can observe from subfigure B of Figure (S6) that our estimates of *R*_0_ are relatively robust with respect to choice of initial values of exposed. The only substantial variation is observed in the first time period - 1^*st*^ - 14^*th*^ April. Now, let us look at how the estimates of number of active cases change with different initial values.

We can observe from subfigure A of Figure (S6) that all the estimates for total active cases increases with increasing values of *E*_0_. The estimate of total active cases on 30^th^ June for *E*_0_ = 4(*U*_0_ + *P*_0_ + *F*_0_) was more than 2 times that for *E*_0_ = (*U*_0_ + *P*_0_ + *F*_0_). Hence we observe that though the estimates of total active cases vary substantially with different initial number of Exposed people, the estimates of Basic Reproduction Number are much more robust to such variations. Now we look at the effect of *α*_*U*_ on our estimates.

##### 12.1.1 Effect of *α*_*u*_

In our main analysis we assumed *α*_*U*_ = 0.5. Here, we try 4 different values of *α*_*U*_, *α* = 0.3, 0.5, 0.7 and 1. First, we look at the estimates of R_0_. Similar to the previous section, from subfigure B of (S7), we observe that the estimates of R_0_ are more or less similar for different values of *α*_*U*_. Once again, the only R_0_ that substantially varies with different values of *α*_*U*_ is the first one i.e. R_01_. Now, we look at the estimates of total active cases.

**Figure S7:**
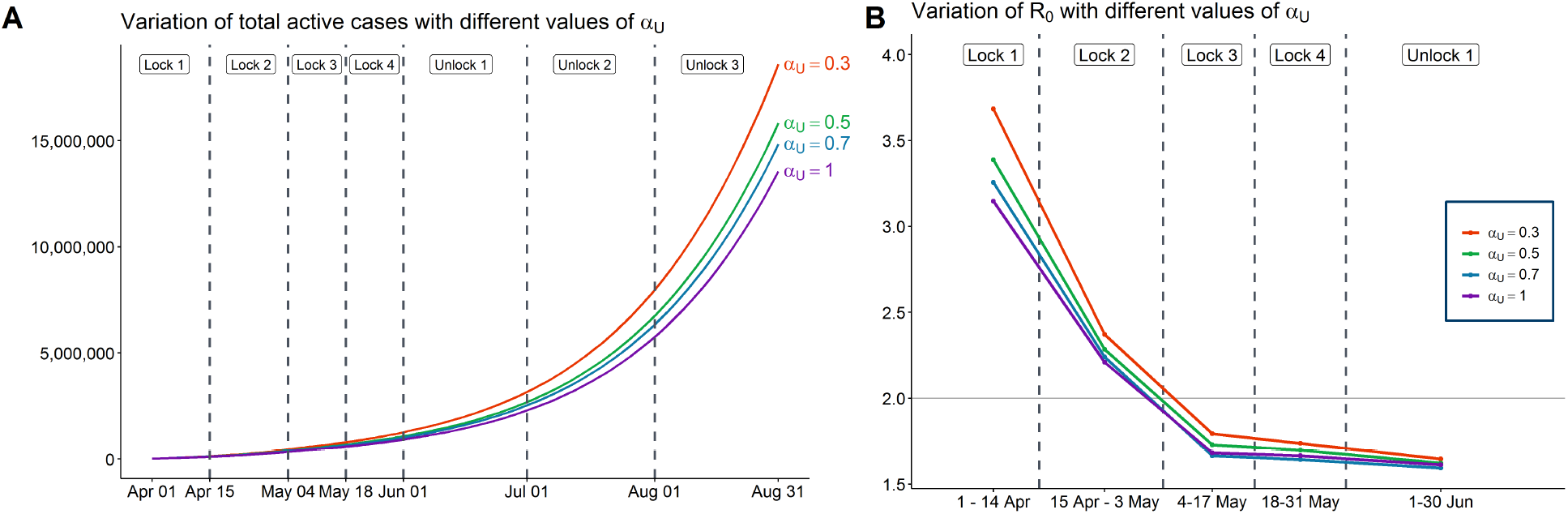
Variation of estimates of *R*_0_ with different values of *α*_*U*_

Subfigure A of (S7) shows that the estimated value of total active cases decreases with increasing value of *α*_*U*_. The reason behind this is if the value of *α*_*U*_ is higher, then a smaller number of untested cases will spread the same amount of infection as a larger number of cases would have if the value of *α*_*U*_ had been lower.

So, once again, we observe that while the estimates of the number of active cases are influenced heavily by *α*_*U*_, the estimates of *R*_0_ remain relatively unaffected by the change.

#### 12.2 Effect of *D*_*e*_

For our main analysis, we had assumed *D*_*e*_ = 5.2. Here, we try 2 more values of *D*_*e*_ and check how our estimates vary with different values of *D*_*e*_.

Again, we observe from subfigure B of (S8), that estimates of *R*_0_ are very robust with respect to different values of *D*_*e*_. From subfigure A of Figure (S8), we note that the predicted number of active cases vary with the different values of *D*_*e*_. However, unlike the previous cases, we do not substantial variation with different variation of *D*_*e*_. Now, we move on to our last sensitivity analysis which is for the value of k in multinomial symptoms.

#### 12.3 Effect of *k*

In Multinomial symptoms model, we defined *k* as the ratio of the probability of a mildly symptomatic individual getting tested to the same for an asymptomatic individual. We chose the value *k* =4 for our main analysis. We had argued why the value of k should be greater than 1 but could not provide any justification for choosing that particular value. So, we try 4 different values of *k* : *k* = 3, 4, 5 and 6. We will start with the estimates of *R*_0_.

Figure subfigure B (S9) shows that similar to previous cases, the estimates of *R*_0_ do not vary much with different values of *k*. We now look at the estimates of total active cases. From subfigure A of Figure (S9), we note that the estimates of total active cases vary with different values of k and with higher values of k we have lower predictions of number of total active cases.

**Figure S8:**
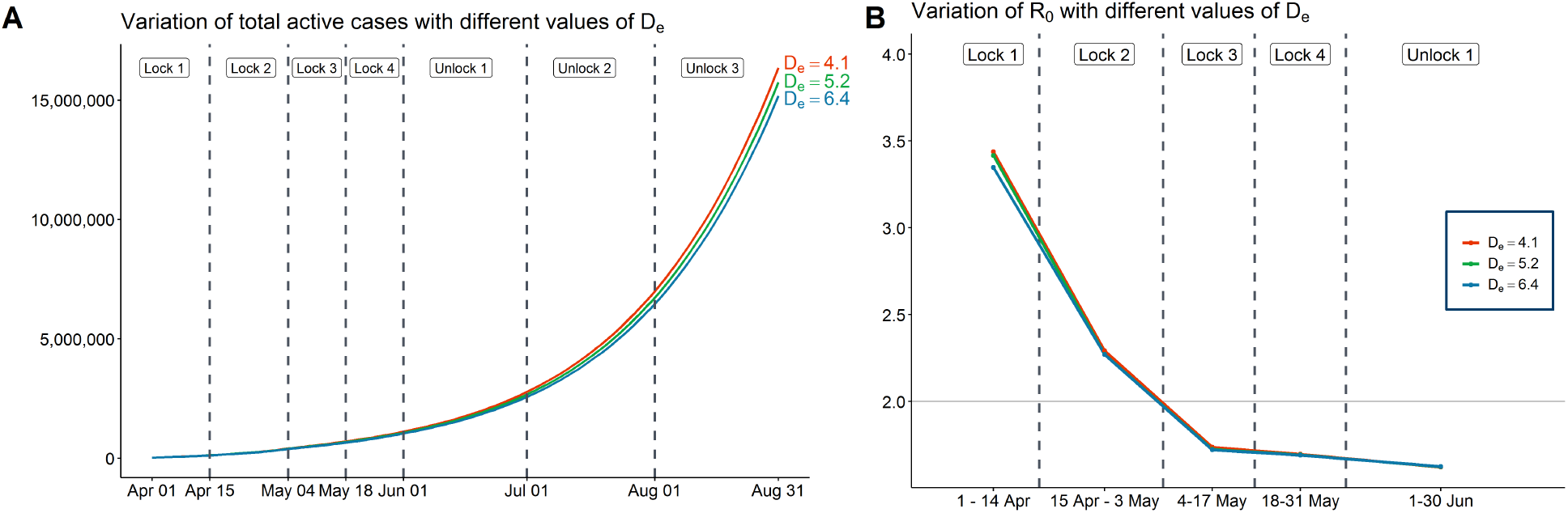
Variation of estimates of *R*_0_ with different values of *D*_*e*_

**Figure S9:**
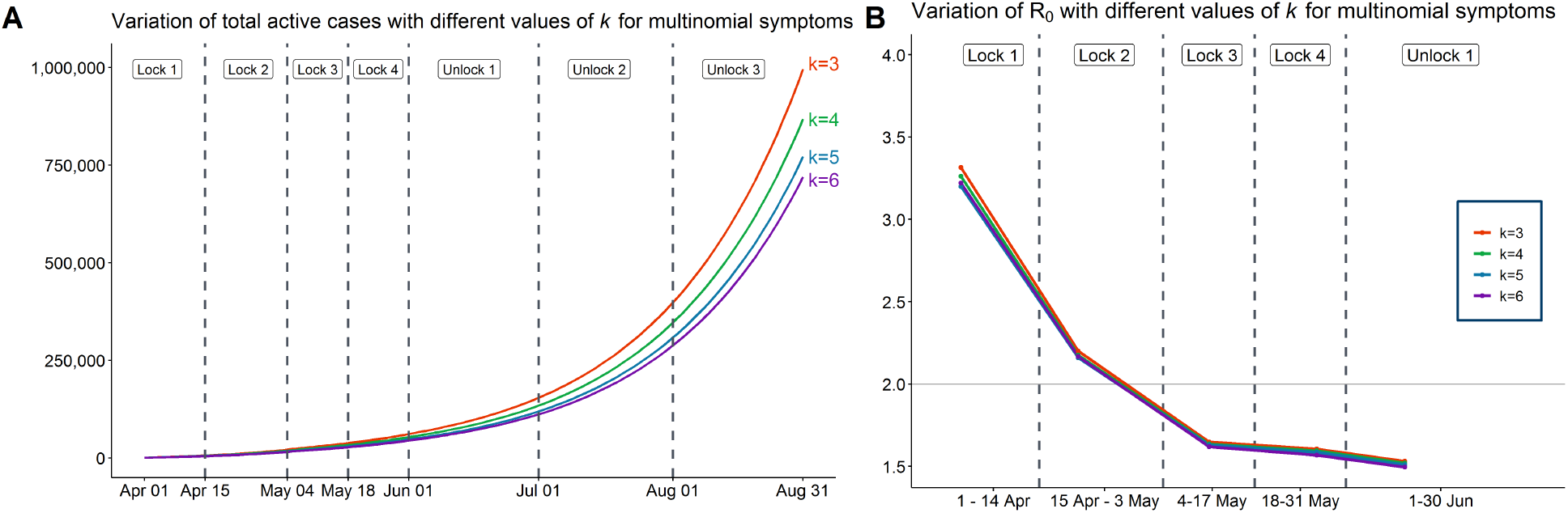
Variation of estimates of *R*_0_ with different values of *k* for multinomial symptoms model

To summarize, we observe that the estimates of the Basic Reproduction number are not substantially influenced by these parameters with an exception of the first Reproduction number. We also note that the estimated number of active cases varies with different values of parameters in most of the cases. It is clearly visible that the sensitivity of the total active case predictions vary across parameters. While it does not vary much with *D*_*e*_, there is substantial variation with different values of *E*_0_.

